# Fibroblast transcriptomics in molecular diagnostics of a comprehensive dystonia cohort

**DOI:** 10.1101/2025.11.14.25339254

**Authors:** Alice Saparov, Ivana Dzinovic, Theresa Brunet, Vicente A. Yépez, Florian Hölzlwimmer, Elisabetta Indelicato, Birgit Assmann, Susann Badmann, Diana Ballhausen, Steffen Berweck, Felix Brechtmann, Melanie Brugger, Kevork Derderian, Felix Distelmaier, Philip Harrer, Denisa Harvanova, Petra Havrankova, Ann-Kathrin Jaroszynski, Miriam Kolnikova, Robert Kopajtich, Anne Koy, Magdalena Krygier, Lukas Kunc, Katarina Kusikova, Oliver Maier, Maria Mazurkiewicz-Bełdzińska, Christian Mertes, Ava Oberlack, Timo Roser, Alexandra Sitzberger, Ugo Sorrentino, Antonia M. Stehr, Katharina Vill, Matias Wagner, Holger Prokisch, Sylvia Boesch, Jan Necpal, Robert Jech, Juliane Winkelmann, Elisabeth Graf, Julien Gagneur, Matej Skorvanek, Michael Zech

**Author notes:** These authors contributed equally to this work (Alice Saparov, Ivana Dzinovic). **Corresponding author:** PD Dr. Michael Zech.

## Abstract

Exome and genome sequencing leave >50% of dystonia-affected individuals without a molecular diagnosis. Where DNA-oriented approaches remain insufficient, integrating multiomics methods and bioinformatics is essential to advance genome interpretation. Herein, we incorporated RNA sequencing (RNA-seq) from a collection of 167 fibroblast samples from individuals affected with dystonic diseases. We leveraged an RNA-seq analysis pipeline, focused on the identification of expression and splicing aberrations, on RNA-seq from skin biopsies. We evaluated a “variant-positive” group of patient samples with preexisting information on variants (36/167, 21.6%), and a “variant-negative” group in which genomic sequencing alone had been unsuccessful in yielding a diagnostic candidate (78.4%). We found that at least 80% of dystonia-associated genes from databases were sufficiently detected by RNA-seq in fibroblasts, highlighting broad applicability. Expression and splicing aberration analyses then produced a manageable number of statistically significant RNA defects affecting dystonia-associated genes for effective case-by-case review. Our approach successfully detected RNA underexpression and mis-splicing for different types of pre-identified dystonia-related variants, providing both benchmarks and insights into mutational mechanisms. Applied to 131 samples from patients without candidate variants from exome and genome sequencing, RNA-seq aided the identification of previously unprioritized causative intronic alterations on reanalysis, providing an added diagnostic yield of 6.9% (9/131). For observed events, we also report the integration of new machine-learning scores predicting corresponding aberrant gene expression in the brain. Fibroblast-based RNA-seq in our selected cohort improved variant interpretation and enabled diagnoses missed by genomic analysis alone, suggesting this framework could be generalized to other dystonias.

## Introduction

Although variants in over 300 genes are linked to dystonic syndromes, diagnostic difficulty and misdiagnoses are still common, undermining the benefits of precise molecular characterization^1^. Intending to resolve a range of undiagnosed presentations of dystonia, we previously created a collaborative research infrastructure that enables prospective application of emerging innovative approaches for etiological discovery^2, 3^. Whole-genome sequencing (WGS) has recently been performed in addition to previous whole-exome sequencing (WES) on difficult-to-diagnose cases from our cohort, but only achieved an added diagnostic yield of 12%^3^. A substantial percentage of WGS variants carried by our study participants were uninterpretable due to indeterminate consequences, resulting in a lack of reporting or prioritization^3^. Despite the introduction of multi-omic workflows for improved DNA-level testing^4–6^, the role of most techniques developed for bi-(or multi-) layered variant analyses is not well defined in the context of dystonia^7^. RNA sequencing (RNA-seq) is a powerful complementary assay for investigating the coding and non-coding genome, allowing for transcriptome-wide detection of aberrant gene expression and defective splicing as evidence for variant pathogenicity as well as specific pathophysiological outcomes^8–10^. A few disease scenarios demonstrated the value of RNA-seq for the evaluation of transcriptional perturbations underlying monogenic conditions based on skin biopsies^11–16^, an easily obtainable source material^9, 11^. However, specific tissue expression profiles of dystonia-linked genes suitable for clinical RNA-seq-based diagnostics have not been thoroughly delineated. Moreover, while RNA-seq has added critical insights for prioritizing pathogenic variants in individual dystonia subjects^3^, systematic diagnostic utilization of this method in a larger patient group has not yet been realized.

Here, we applied an automated RNA-seq analysis pipeline^17^ to dermal fibroblast lines from 167 unrelated dystonia-affected individuals, the largest compendium collected in this indication to date. Defining a catalog of detectable dystonia-related genes in fibroblast RNA-seq samples, we begin characterizing the effects of different dystonia-linked variant types on RNA phenotypes. In patients where DNA-sequencing data alone provided no diagnosis, comprehensive, computationally-driven investigation of aberrant RNA-expression and/or splicing events could help in downstream causative variant identification. Since neuronal cell expression was not measured, we additionally explored machine learning model-derived predictions of aberrant RNA-underexpression^18^ associated with discovered variants in dystonia-relevant brain regions.

## Methods

### Dystonia fibroblast collection

Using our multicenter network^2, 3^, fibroblast samples were collected from 167 index patients (77 female, 46.1%; mean age at sampling 21.0 years, range 1-69 years) who were clinically diagnosed with rare dystonic syndromes^19^ (**Fig.1**). These patients underwent standard punch biopsies for skin-specimen acquisition by collaborators in Austria, Czechia, Germany, Poland, Slovakia, and Switzerland. Informed written consent was obtained, the institutional ethics review boards of the collaborating partners approved the procedures, and the study was conducted in accordance with the principles of the Declaration of Helsinki. The set of enrolled individuals had extensive prior genomic investigations, including WES-only (13 cases with likely pathogenic/pathogenic^20^ or candidate variants from this analysis), and WES with follow-up WGS studies (*n* = 154)^2, 3^. We favored both cases with pre-existing findings from WES/WGS (“variant-positive” group) and cases without prioritized variants but with suspicion of an underlying genetic disorder^3, 21^ (“variant-negative” group) for fibroblast-culture establishment and RNA-seq (**Fig.1**): pre-identified variants of interest, often affecting newly discovered or less-established dystonia-related genes, were present in 36 individuals (21.6%); in 16 of the subjects (16/36, 44.4%), RNA-seq provided additional evidence (*n* = 14) or was required (*n* = 2) for classifying these variants as likely pathogenic/pathogenic^20, 22^ (**Fig.1**; see Results and **Suppl.Tab.1**); the remaining 131 patients (78.4%) lacked a selected variant candidate despite comprehensive screening of WES/WGS outcomes for multiple mutation types^3^ at the time of skin-sample collection and were enrolled for RNA-seq data-aided genomic reanalysis^14, 23^. The majority of patients with sampled fibroblasts displayed early-onset (85.0%), non-focal (92.2%), and/or combined dystonia (76.0%); a summary is given in **Fig.1**. The control fibroblast-based RNA-seq cohort included samples from 182 unrelated research donors who had been referred to our sequencing platform for various clinical indications except dystonia. **The patient IDs mentioned in this work were not known to anyone outside the research group.**

**Table 1.** Index patients with new diagnostic findings after genomic reanalysis aided by RNA-seq.

| Index patient<sup>3</sup> | Phenotype | Gene/ diagnosis (OMIM) | Variant(s) | Variant type/ zygosity | Detected significant outlier event(s); RNA phenotype |
| --- | --- | --- | --- | --- | --- |
| **Extended splice-region variants ($\pm 3\text{bp}$ to $\pm 10\text{bp}$ region)** |  |  |  |  |  |
| R030 | dystonia, spasticity, ataxia | *ACP33 (SPG21)/*<br>Mast syndrome (248900) | NM_016630.7: c.306+6T>A | intronic SNV/ hom | *ACP33 (SPG21)*: AS; exon-4 skipping<sup>1</sup> |
| R054 | dystonia, ataxia, ID, gingival hyperplasia, cerebellar hypoplasia | *ATG7/*<br>spinocerebellar ataxia-31 (619422) | NM_001349232.2: c.528+3A>G/<br>c.1090G>A, p.Gly364Ser | intronic SNV/ missense<br>SNV/ comp het | *ATG7*: AS; exon-8 skipping, partial retention of introns 7 and 8 |
| R089 | dystonia, DD, epilepsy | *GLS/*<br>developmental and epileptic encephalopathy-71 (618328) | NM_014905.5: c.1713-9C>G | intronic SNV/ hom | *GLS*: AS; exon-16 extension, skipping of exons 16 and 17<sup>1</sup> |
| **Deep(er) intronic variants ($> \pm 10\text{bp}$ from exon-intron boundaries)** |  |  |  |  |  |
| R082 | dystonia, chorea, ID | *SHQ1/*<br>neurodevelopmental disorder with dystonia and seizures (619922) | NM_018130.3: c.144-24A>G/<br>c.487G>T, p.Asp163Tyr | intronic SNV/ missense<br>SNV/ comp het | *SHQ1*: AE, AS; significant RNA underexpression (FC: 0.51), exon-2 extension (c.144-24A>G), exon-5 skipping (c.487G>T) |
| R140 | dystonia, stereotypies, DD, cerebellar atrophy | *SNX14/*<br>spinocerebellar ataxia-20 (616354) | NM_153816.6: c.1108+28A>G | intronic SNV/ hom | *SNX14*: AE, AS; significant RNA underexpression (FC: 0.23), exon-12 extension |
| R158 | dystonia, DD, eye movement abnormalities | *AGTPBP1/*<br>neurodegeneration, childhood-onset, with cerebellar atrophy (618276) | NM_001330701.2: c.1303-4238C>G | intronic SNV/ hom | *AGTPBP1*: AE, AS; significant RNA underexpression (FC: 0.20), pseudoexon splicing-in |
| R134<sup>2</sup> | dystonia | *ATM/*<br>ataxia-telangiectasia (208900) | NM_000051.3: c. 3284+695G>T;<br>c.3284+699A>C | intronic MNV/ hom | *ATM*: AE, AS; significant RNA underexpression (FC: 0.38), pseudoexon splicing-in<sup>1</sup> |
| R105<sup>2</sup> | dystonia,<br>spasticity | *SPG11*/<br>spastic paraplegia-11 (604360) | NM_025137.4: c.1235C>G,<br>p.Ser412*/ c.3454-28A>G | nonsense SNV/ intronic<br>SNV/ comp het | *SPG11*: AE, AS; significant<br>RNA underexpression (FC:<br>0.22), exon-20 skipping, partial<br>retention of introns 19 and 20<sup>1</sup> |
| R058<sup>2</sup> | dystonia,<br>spasticity,<br>DD | *UFC1*/<br>neurodevelopmental disorder with spasticity and poor<br>growth (618076) | NM_016406.4: c.244_255del,<br>p.Glu83_Ile86del/ c.255+17G>A | inframe indel/ intronic<br>SNV/ comp het | *UFC1*: AE, AS; significant RNA<br>underexpression (FC: 0.59),<br>exon-3 skipping<sup>1</sup> |

<sup>1</sup>Significant underexpression of the encoded protein detected on proteomics analysis (proteomics data available for fibroblast samples from patients R030, R089, R134, R105, and R058).
<sup>2</sup>Index patient and causative intronic variants previously reported in detail in a multi-omics companion manuscript (PMID: 39937650).
<sup>3</sup>**The patient IDs were not known to anyone outside the research group.**
Abbreviations: AE, aberrant expression; AS, aberrant splicing; bp, base pairs; comp het, compound heterozygous; DD, developmental delay; F, female; FC, fold change; hom, homozygous; ID, intellectual disability; M, male; MNV, multi-nucleotide variation; OMIM, Online Mendelian Inheritance in Man; RNA-seq, RNA sequencing; SNV, single-nucleotide variant; y, years.

**Figure 1.**
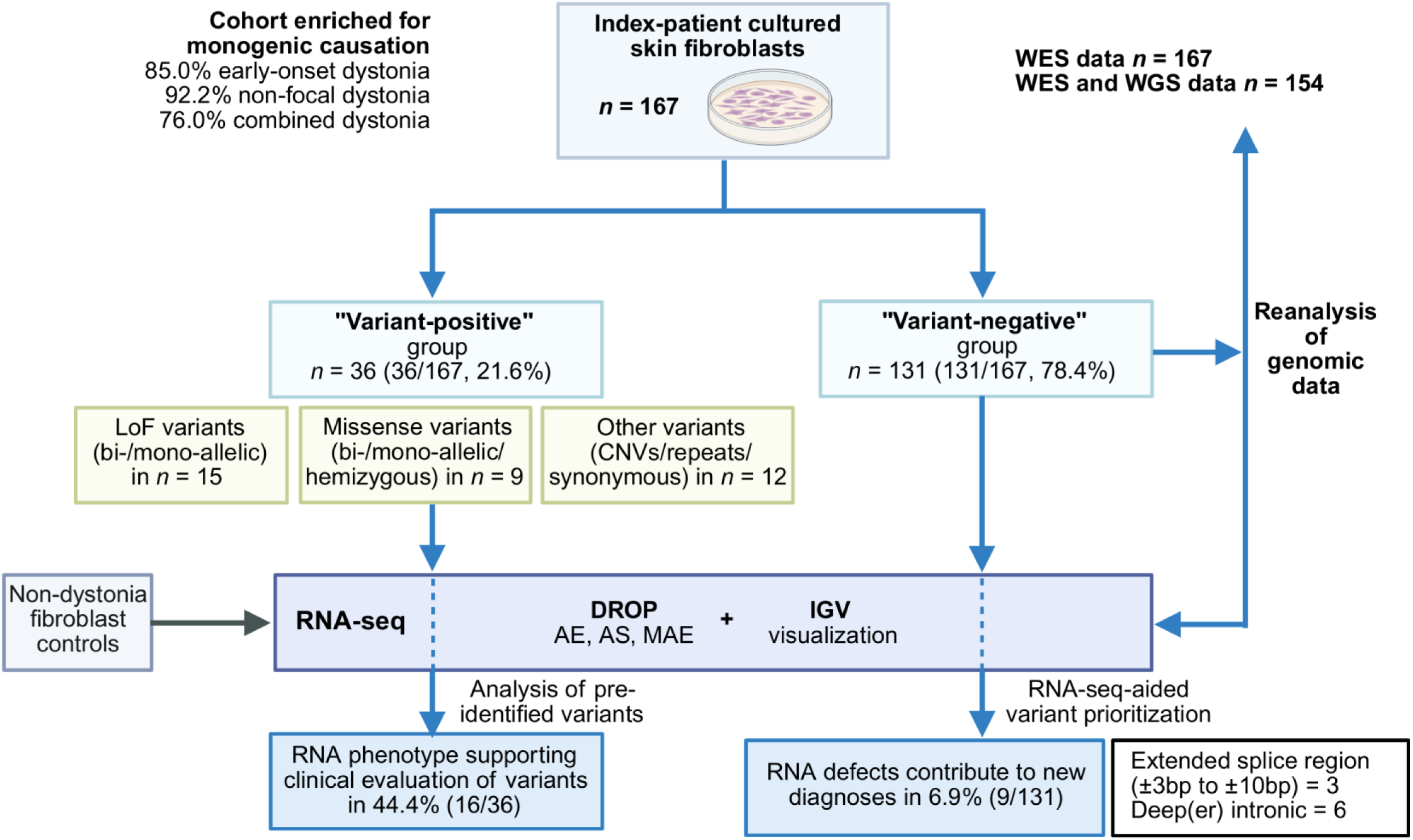
Overview of the RNA-seq pipeline for fibroblasts from dystonia-affected individuals. All recruited skin fibroblast lines from patients with pre-identified variants (“variant-positive” group, 21.6%) and patients for whom WES/WGS alone had not yielded a relevant diagnostic variant candidate (“variant-negative” group, 78.4%) received RNA-seq. The data were analyzed using the DROP workflow^17^, direct visualization in IGV^24^, and comprehensive genomic reevaluation. We assessed the outcomes of RNA-seq for both patient groups, considering the range of aberrant RNA phenotypes resulting from various variant categories, the benefits in aiding clinical interpretation, and the added diagnostic yield. AE, aberrant expression; AS, aberrant splicing; CNVs, copy-number variants; DROP, Detection of RNA Outliers Pipeline; IGV, Integrative Genomics Viewer; LoF, loss-of-function; MAE, mono-allelic expression; RNA-seq, RNA sequencing; WES, whole-exome sequencing; WGS, whole-genome sequencing.

### RNA-seq

RNA sequencing and processing for dystonia cases and controls were performed according to standard methodologies, integrating established equipment and reagents, protocols, and quality-control parameters^3, 11, 14^. RNA libraries were created using Illumina Stranded mRNA Prep Ligation, polyA-tailed kits, ensuring consistency of the samples with our previous RNA-seq projects^14^. A NovaSeq6000 instrument (Illumina) was used to produce 100-bp paired-end reads with a mean number of 76.7 million (range: 44.9-146.6 million) read pairs per sample. Downstream analysis of raw data was carried out with an earlier described bioinformatics workflow^14, 17^. STAR v.2.4.2a was deployed to align reads to the GRCh37/hg19 reference with high mapping accuracy; on average, 85.7% of uniquely mapped reads were achieved for all samples. The processed alignment files were used for visualization of genomic variant-related RNA defects in the Integrative Genomics Viewer (IGV)^24^, and as input for parallelized analysis of aberrant events in the Detection of RNA Outliers Pipeline (DROP)^17^, comprising our standardized framework for transcriptome-wide studies.

### RNA expression profiles and disease-gene lists

DROP output was used to determine the dystonia genes expressed in fibroblasts^17, 23^. “Expressed genes” were defined in DROP as genes for which >5% of the analyzed samples exhibited Fragments Per Kilobase of transcript per Million mapped reads (FPKM) values of >1. These expressed genes were filtered with disease-gene lists, which were obtained as follows:

(i) Online Mendelian Inheritance in Man (OMIM)^25^ and published compendia^26^ were queried for human morbid genes^25^ and genes associated with dystonia^25^ or “rare neurological disease”^26^;
(ii) MDSGene^27^ and GeneReviews^28^ were accessed to extract curated descriptions of isolated dystonia-related genes; and (iii) dystonia-pathway genes were manually reviewed as an expansion of a compendium recently compiled by us^29^. To compare the results from RNA-expression profiling in fibroblasts to a second clinically accessible tissue, we also examined the numbers of expressed dystonia-related genes in 279 whole-blood RNA-seq samples that were available to us from a different research project.

### Global RNA-defect studies

Evaluating outlier status for expression, splicing, and allele imbalance in RNA-seq data involved previously reported procedures^17^, streamlined on our locally installed bioinformatics infrastructure^14^. We employed DROP^17^ v.1.4.0 (the latest at the time of analysis), incorporating the denoising autoencoder-based algorithms Outlier in RNA-Seq Finder (OUTRIDER)^30^ v.1.20.1 for identification of aberrant expression (AE) and Find RAre Splicing Events in RNA-seq (FRASER) 2.0^31^ v.1.99.4 for discovery of aberrant splicing (AS). A false-discovery rate (FDR) threshold of ≤0.05 after multiple-testing correction was used for significant AE outcomes, whereas an FDR of ≤0.05 with an effect size |*Δ*J| ≥0.1 was required for significant AS events. We also ran a negative binomial test-based tool for the detection of mono-allelic expression (MAE), as introduced by Kremer et al.^11, 17^. Subsequently, expression abnormality-associated *z*-score and fold-change values, as well as splice-defect metrics and MAE hits were explored at an individual patient basis. Considering the expected rarity of individual causative molecular lesions, our analytic strategies were based on the comparison of one case subject against all the remaining samples (dystonia patients and controls, *n* = 349)^14^.

### Association of pre-identified variants with RNA phenotypes

For each patient from the “variant-positive” group, we assessed potential RNA aberrations (AE, AS, MAE) in the context of previous genomic findings. Genes and variants of interest were associated with the corresponding outcomes from DROP^14^. We also analyzed rankings that incorporated expression information from all patient and control samples^3, 32^. The pre-identified variants were further manually investigated for possible effects on splicing patterns or associated allele-biased expression using simultaneous visualization of RNA-seq and WES/WGS data in IGV.

### Reanalysis of WES/WGS variants aided by RNA-seq data

DROP results for AE, AS, and MAE were used in the case-by-case reanalysis of WES/WGS data in the “variant-negative” group^14^. For clinically meaningful evaluation, we narrowed the transcriptome-wide outlier summary files down to significant hits (FDR≤ 0.05) that were associated with monogenic disorders in OMIM and specifically focused on genes linked to dystonia and related neurological conditions consistent with the observed phenotypic presentations^25^. MAE analysis was restricted to hits associated with rare variants (minor-allele frequency <0.001 in gnomAD and <0.05 in dystonia-patient and control datasets; allelic imbalance >85%). Informed by the DROP-output shortlists, we revisited variants affecting plausible genes in the individual patients; expanded search strategies for previously neglected types of coding and non-coding rare variations that matched the expected zygosity for the disease in question were implemented. Resulting priority events were visually inspected in IGV at the RNA and DNA levels. All combined interpretations of RNA defects, WES/WGS variants, and disease phenotypes were undertaken by genetics specialists and discussed with referring clinicians to reach consensus on diagnosis^10, 14, 20^. Results from complementary tests were taken into account when possible, including findings from additional segregation, proteomics^3^ (**Tab.1**), laboratory, and reverse-phenotyping analyses.

### Prediction of variant-related aberrant RNA expression in the brain

We modeled the impact of patient-specific genetic perturbations on RNA expression in dystonia-relevant brain regions using the recently established algorithm AbExp^18^. Identified variants that were associated with corresponding significant gene-underexpression outliers in fibroblasts were included in the analysis.

## Results

### Dystonia-related genes covered by fibroblast-based RNA-seq

To demonstrate skin fibroblast utilization as an appropriate sample type for broader RNA-seq-based diagnostic testing in dystonias, we first compiled lists of dystonia-related genes and assessed their expression patterns using a recommended threshold of >1 FPKM^17, 23^. Consistent with previous results^14, 15^, an average of 15,180 total genes were detected through RNA-seq in fibroblasts from our patients. Overall, the fibroblasts expressed 80.5% (306/380) of genes linked to dystonic disorders in OMIM^25^ (**Fig.2A**). This corresponded to a higher percentage of testable genes compared with the expression of all OMIM-morbid genes (66.1%, 3109/4707)^25^, and a similar rate of gene coverage compared with a comprehensive, recently published “rare neurological disease gene” set (79.6%, 1448/1820)^26^ (**Fig.2A**). When focusing on genes primarily involved in isolated dystonia according to reference databases (*n* = 10)^27, 28^, we observed consistently high coverage (80%); only *GNAL* and *HPCA* did not reach the expressed gene threshold (**Fig.2B**). Gene-informed molecular pathways are increasingly recognized in dystonia and provide insights into pathogenic mechanisms^1^. Thus, we investigated what fractions of genes within biologically defined pathways for dystonic conditions^29^ could be analyzed by RNA-seq. Our fibroblast-based approach detected expression of all genes in 1 of 10 pathways, whereas measurable dystonia-related gene expression profiles between 43% and 90% were found for the remaining 9 pathways (**Fig.2C**). Most pathways had detectable gene representations of >60%-80% (**Fig.2C**). As the discovery of gene underexpression caused by loss-of-function (LoF) variants was a major contributor to diagnostic success in previous RNA-seq studies^12–14^, we also explored how many dystonia-related pathway genes were associated with LoF mechanisms; of those pathway genes expressed in fibroblasts, ∼82.1% (165/201) were implicated in haploinsufficiency, X-linked, or recessive disorders with reported causative null alleles^25, 33, 34^, indicating that fibroblasts represented a suitable tissue for RNA-seq-based evaluation of a variety of dystonia-related genes that participate in key cellular disease processes. Finally, comparison of gene expression between blood and fibroblasts highlighted that the latter provided a more comprehensive transcriptional landscape for successful application of RNA-seq-based dystonia diagnostics (only 71.8% [273/380] of dystonia-related genes from OMIM expressed in blood, **Fig.2A**). Our expression benchmark illustrated the effectiveness of the RNA-seq analysis pipeline based on fibroblast cell lines in identifying many relevant dystonia-related genes, establishing a compendium of testable targets for RNA-driven diagnostic strategies.

**Figure 2.**
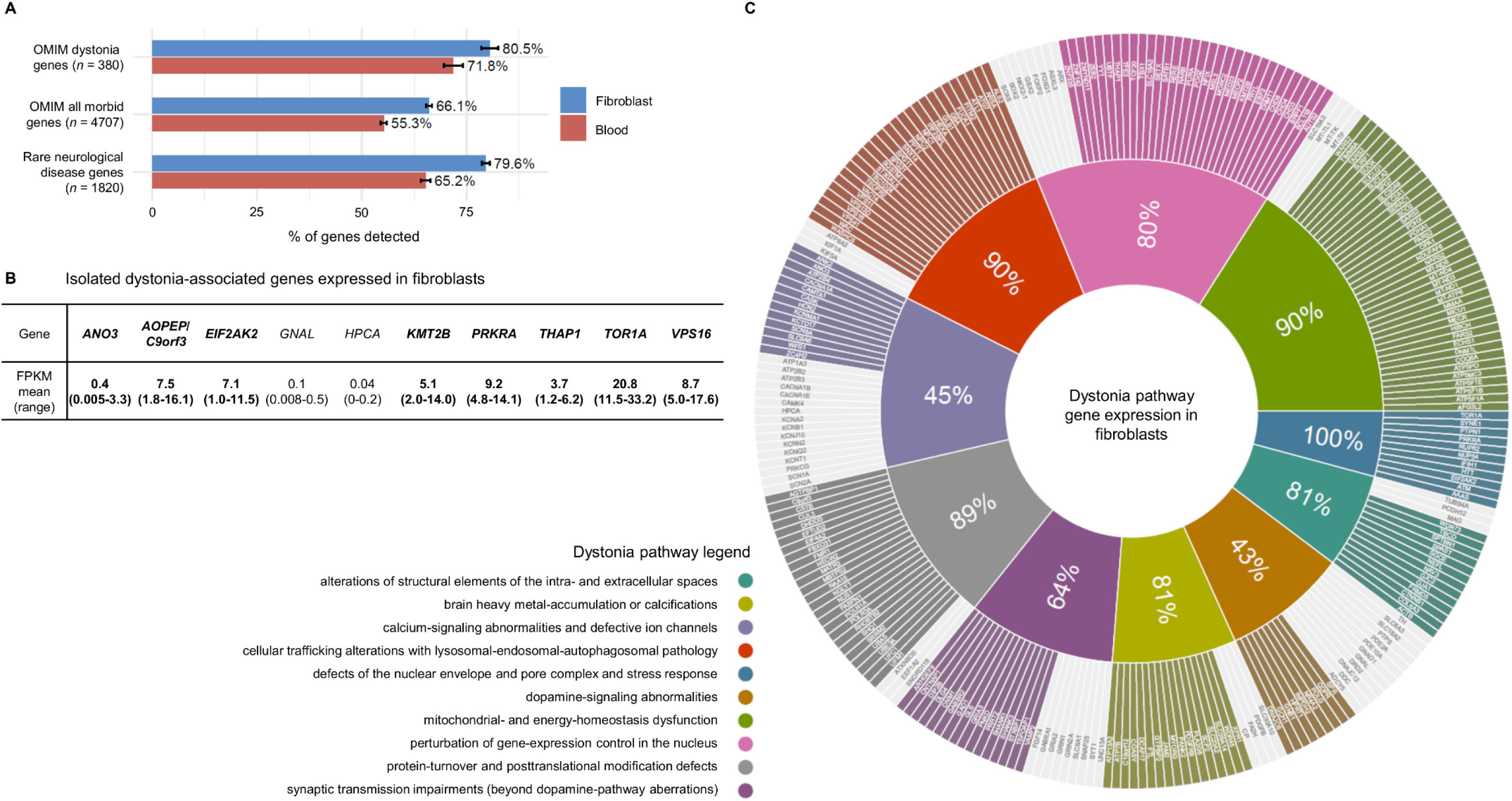
Detection of dystonia-associated gene expression by RNA-seq. (**A**) Percentages of different gene sets that were covered by RNA-seq, as defined by the DROP workflow^17^, in cultured skin fibroblasts from our patients and controls (*n* = 349). Dystonia-associated genes (*n* = 380) and all human morbid genes (*n* = 4707) were obtained by systematic OMIM queries^25^, as described^2, 3^. The “rare neurological disease gene” set (*n* = 1820) has been published recently^26^. Corresponding gene coverages in blood samples are provided for comparison. For each percentage of detectable genes, errors bars represent standard errors of the mean. (**B**) Summary of curated isolated dystonia-associated genes^27, 28^ and their detectable expression in all fibroblast samples included in this study. All genes except for two (*HPCA*, *GNAL*, not shown in bold) reached the “expressed gene” threshold^17, 23^. FPKM scores (mean, ranges) in our 349 total samples are given for each gene. (**C**) Dystonia-associated gene coverage by RNA-seq according to 10 reported disease-relevant molecular pathways^29^. Percentages of covered genes are highlighted. The pathway genes were manually curated based on previous reviews^1, 29^ and continuous searches of publicly available data^25^, but the lists cannot be regarded as exhaustive complete collections and it is possible that other relevant genes were not added. Despite this limitation, the given percentages are likely representative of the approximate fractions of genes that are covered by RNA-seq in the different pathways. Color code is provided for the pathways. DROP, Detection of RNA Outliers Pipeline; FPKM, Fragments Per Kilobase of transcript per Million mapped reads; OMIM, Online Mendelian Inheritance in Man; RNA-seq, RNA sequencing.

### Aberrant events detected by automated RNA-seq data analysis

We used DROP to assess RNA defects transcriptome-wide in patient fibroblasts^14, 17^, namely AE, AS, and MAE^17^; results are summarized in **Fig.3A-C**. OUTRIDER uncovered on average 5 significant expression outliers (range: 0-39) per patient. Underexpression was more commonly observed, with 72.9% (643/882) of all significant outliers in patients demonstrating fold changes (FCs) between 0.01 and 0.84. In the FRASER 2.0 analysis with standard settings, each patient sample typically presented 7 splicing outliers (mean; range: 0-27). The mean number of rare variants with significant allelic imbalance per sample was 3 (range: 0-19). When refining the analysis by only considering dystonia-related genes according to OMIM^25^, a range of 0-2, 0-3, and 0-1 hits were identified in each of the patients by OUTRIDER, FRASER 2.0, and the MAE-detection module, respectively. Collectively, our RNA-seq analysis pipeline yielded a manageable number of filtered DROP results, enabling effective manual evaluation of the associations between genomic variants and transcriptional aberrations.

**Figure 3.**
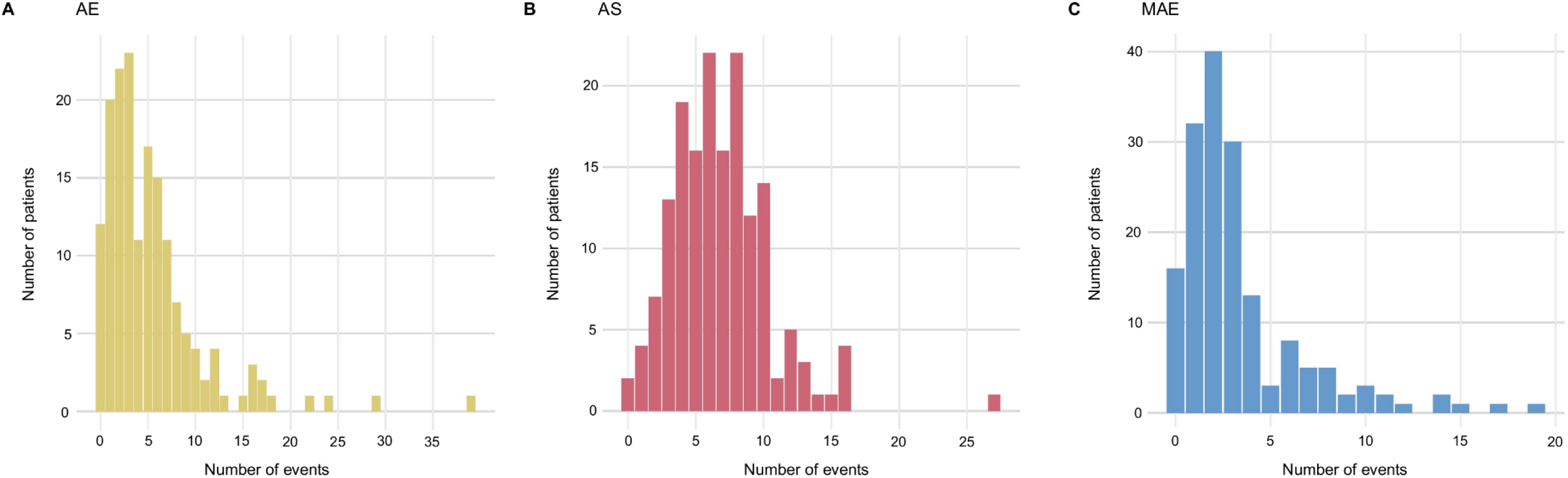
Summary of significant outlier events identified by automated bioinformatic analyses in dystonia RNA-seq samples. We applied the DROP workflow^17^ to the RNA-seq data from our 167 dystonia-affected individuals, allowing for systematic identification of AE, AS, and MAE events through the OUTRIDER^30^, FRASER 2.0^31^, and allele-biased expression discovery^11, 14^ modules, respectively. Distributions of detected significant AE hits (FDR ≤ 0.05; *n* = 882 total events) (**A**), significant AS hits (FDR ≤ 0.05; *n* = 1122 total events) (**B**), and significant MAE hits (FDR ≤ 0.05; *n* = 553 total events) (**C**) for the patient cohort are shown. Only rare genomic variants (MAF <0.001 in gnomAD and <0.05 in the present patient and control cohort) were considered in the MAE analysis (allelic imbalance >85%). AE, aberrant expression; AS, aberrant splicing; FDR, false-discovery rate; FRASER, Find RAre Splicing Events in RNA-seq; MAF, minor allele frequency; MAE, mono-allelic expression; OUTRIDER, Outlier in RNA-Seq Finder; RNA-seq, RNA sequencing.

### RNA abnormalities in patients with pre-identified variants

Investigating the additive value of RNA-seq data on different pre-identified dystonia-associated variants was evaluated through transcriptional outcomes and enhancements to their clinical interpretations. Of the 36 “variant-positive” samples included in this evaluation (27 [75.0%] of which were previously published in the frame of single-gene or larger cohort studies^3, 32, 35–39^; **Suppl.Tab.1**), 15 carried bi-allelic (*n* = 2) or mono-allelic (*n* = 13) likely pathogenic/pathogenic^20^ LoF variants; 9 had bi-allelic (*n* = 2) or mono-allelic/hemizygous (*n* = 7) missense variants (likely pathogenic/pathogenic^20^ variants in 7 samples; variants of uncertain significance [VUS]^20^ in 2 samples); and 12 were known to harbor other mutation types, either classified as likely pathogenic/pathogenic^20, 40, 41^ alterations (9 samples) or as VUS^20, 40^ (3 samples), including copy-number variants (CNVs), repeat expansions, and synonymous variants (**Suppl.Tab.1**). Examining samples with LoF changes showed that perturbation of both alleles resulted in significant gene underexpression (*POLR3A*, FC: 0.57; *ZNF142*, FC: 0.67; **Fig.4A/B**; **Suppl.Tab.1**), whereas the mono-allelic LoF variants had variable outcomes from our RNA-seq analysis (**Fig.4A/B**; **Suppl.Tab.1**). In 8/13 (61.5%) of the cases with heterozygous LoF variants, we detected significantly decreased expression of the variant-bearing genes, demonstrating that RNA-seq can readily aid in identifying gene-inactivating alleles and in supporting haploinsufficiency as a pathomechanism (**Fig.4A/B**; **Suppl.Tab.1**). OUTRIDER successfully revealed significant AE of *VPS16* (FCs: 0.58-0.59; **Fig.4A/B**; **Suppl.Tab.1**) in 3 unrelated cases with previously diagnosed *VPS16*-related dystonia^3, 35^, a known common form of autosomal dominant isolated dystonia^1^. Clinical significance assessment of LoF variants was also improved in the context of newly described disease genes and novel gene-phenotype correlations. For example, interrogation of AE outliers in patient R083 carrying a heterozygous nonsense substitution in *PTPN1*, a gene that has recently been implicated in dystonic encephalopathy^36^, confirmed significant reduction of *PTPN1* RNA levels (FC: 0.65) and thus the haploinsufficiency effect of the variant (**Fig.4A/B**; **Suppl.Tab.1**). Similarly, we found two genes for which primarily disease-causing missense mutations had been reported^32^, *ATP5F1A* and *ATP5F1B*, to be significantly down-regulated by heterozygous LoF variants (FCs: 0.63 and 0.69, respectively; **Fig.4A/B**; **Suppl.Tab.1**), expanding the genotypic spectrum of mitochondrial ATP synthase-related dystonias^32^. Although OUTRIDER did not detect corresponding significant outliers in RNA-seq data for the remaining likely pathogenic/pathogenic heterozygous LoF alterations (**Fig.4A/B**; **Suppl.Tab.1**), closer analysis of the results using cohortwide rank-based methods^3^ showed that one variant (frameshift variant in *ANK2*^3^) was associated with the lowest expression of the affected gene in comparison to all other samples (sample rank: 1/349, FC: 0.51; OUTRIDER FDR>0.05; **Fig.4B**); in contrast, a frameshift change in the single-exon gene *IRF2BPL*^3^ caused no RNA underexpression (sample rank: 225/349, FC: 1.04; **Fig.4B**), consistent with previous findings from RT-PCR studies of *IRF2BPL* mutations^42^. This emphasized the value of complementary analysis strategies in large RNA-seq datasets to characterize different effects of LoF variants on expression. In line with earlier findings^11, 14^, for most pre-identified bi-allelic and mono-allelic/hemizygous missense variants, we observed no significant RNA outlier events (8/9, 88.9%; **Fig.4A/B**; **Suppl.Tab.1**). However, when present, as seen in patient R072 (**Fig.4C**; **Suppl.Tab.1**), RNA-seq prompted reconsideration of the significance of a predicted missense variant. Trio-WES yielded a *de novo* hemizygous c.970G>A (p.Ala324Thr) substitution in *MBTPS2*, initially classified as a VUS attributed to R072’s presentation, consisting of dystonia, developmental delay, and epilepsy, which did not match the typical phenotype of *MBTPS2*-related IFAP/BRESHECK syndrome^25^; RNA-seq uncovered a *MBTPS2* splicing defect (exon skipping, intron retention) along with significant *MBTPS2* underexpression (FC: 0.65) associated with the variant, thereby suggesting that *MBTPS2* deficiency may well contribute to this patient’s clinical outcome. For 2 pre-identified CNV calls in affected individuals, our RNA analysis provided supporting evidence for these variations (**Fig.4A/B**; **Suppl.Tab.1**), directly identifying dystonia-associated structural variants that can be challenging to detect by DNA-sequencing workflows alone^5^. One example was found in patient R139, where a heterozygous 6q22.1-q22.31 microdeletion encompassing *NUS1*^39^ led to approximately half of the normal expression of 6 adjacent genes located within the CNV (significant outliers in OUTRIDER, FCs: 0.44-0.59; **Fig.4D**; **Suppl.Tab.1**). Our pipeline further allowed for the detection of significant AE of genes affected by autosomal-recessively inherited repeat expansions^3^, including *CSTB* (FC: 0.31) and *GLS* (FC: 0.59), whereas none of the 5 pre-identified repeat expansions associated with autosomal dominant/X-linked inheritance produced a detectable RNA defect. Finally, for 3 cases with synonymous VUS, each observed in *trans* with another variant type (**Fig.4A/B**; **Suppl.Tab.1**), no statistically significant AE status of the affected genes or resultant AS or MAE calls were found by our automated pipeline. However, lowered expression of *HCN2* was evident (sample rank: 1/349, FC: 0.32, OUTRIDER FDR>0.05; **Fig.4B**) in association with a heterozygous, predicted synonymous c.1560C>T substitution and a heterozygous exon-6 deletion in patient R047 with dystonia and epileptic encephalopathy (**Suppl.Tab.1**). Visualized inspection of RNA-seq data indicated that c.1560C>T created a cryptic splice donor, leading to exon-5 truncation (data not shown). While the splicing defect was only evident in a few RNA reads, likely because of nonsense-mediated decay (NMD)^15^, the finding indicated the disruptive nature of c.1560C>T and established the diagnosis of autosomal recessive *HCN2*-related neurodevelopmental disorder; the case exemplified the need for careful variant-guided inspection of RNA-seq data, in parallel to pipeline-based analyses. A total of 3 out of 36 pre-identified variants (8.3%) could not be examined by RNA-seq because the affected genes were not expressed in skin fibroblasts (*ADCY5*, *ATP2B2*, *ATXN8OS/SCA8*; Suppl.Tab.1).

**Figure 4.**
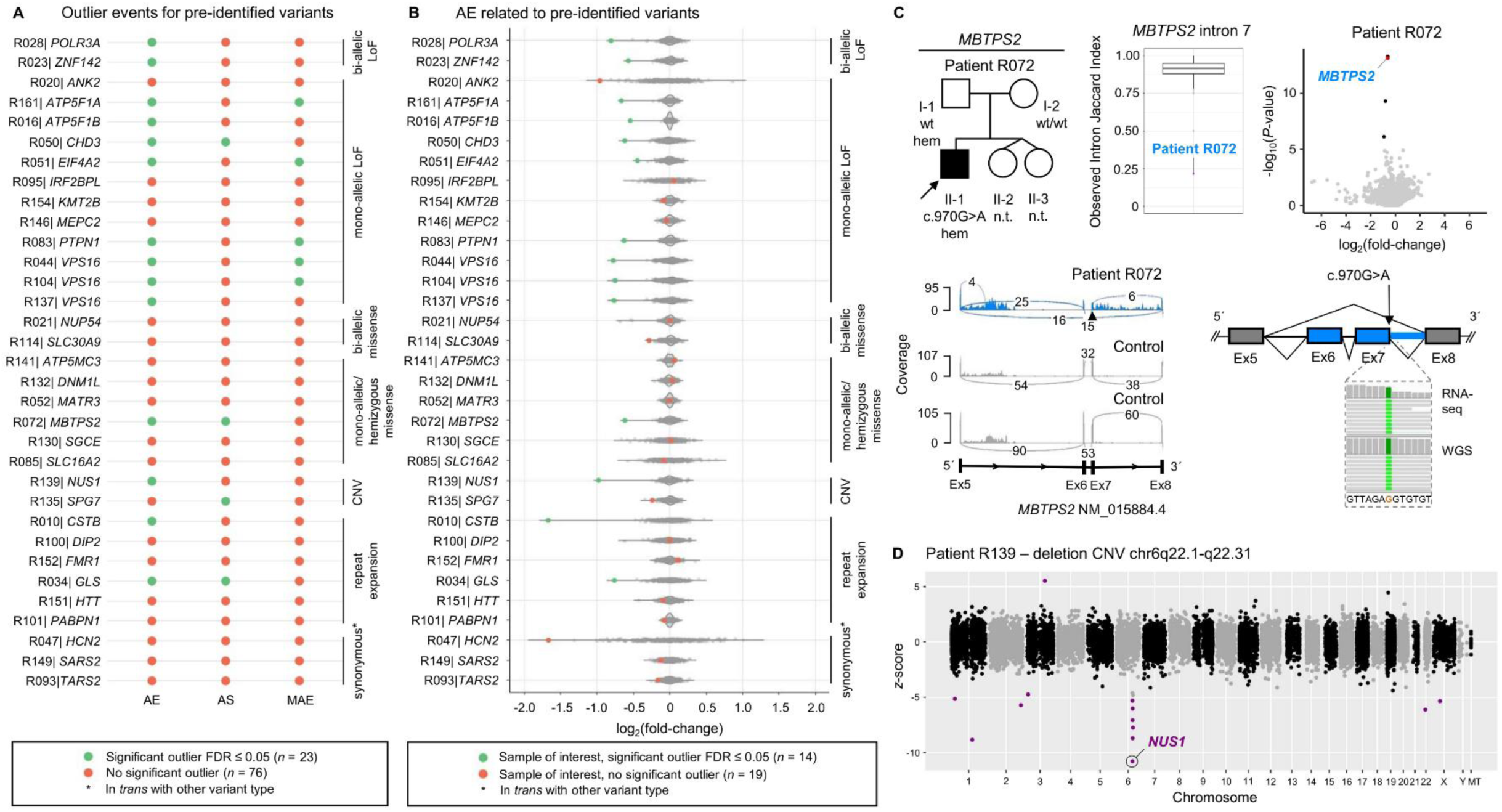
RNA phenotypes associated with pre-identified variants and examples of readouts aiding in clinical interpretation. (**A**) Summary of DROP-workflow^17^ outcomes for all pre-identified variants that were expressed in fibroblasts and assessed by RNA-seq in this study. The colored dots indicate whether or not a given WES/WGS-identified variant produced a significant outlier hit in the AE, AS, and/or MAE analyses. Color code is provided. Samples with variants in *ADCY5*, *ATP2B2*, and *ATXN8OS/SCA8*, unexpressed in fibroblasts, are not shown. (**B**) Depiction of log2fold changes of RNA expression of the gene of interest for each sample with a pre-identified variant (red or green dots) compared to all other samples in which the gene was expressed. Data are based on the output from OUTRIDER^30^, and each dot indicates a sample. Gene-wise outlier events representing statistically significant RNA underexpression (AE with FDR ≤ 0.05) are shown in green; color code is provided. Samples with variants in *ADCY5*, *ATP2B2*, and *ATXN8OS/SCA8*, unexpressed in fibroblasts, are not shown. (**C**) RNA-seq-aided diagnosis of *MBTPS2*-related IFAP/BRESHECK syndrome in patient R072, highlighted by an arrow in the pedigree. OUTRIDER^30^ and FRASER 2.0^31^ identified significant outlier events for *MBTPS2*, as represented in the RNA expression volcano plot (underexpression outlier for *MBTPS2* highlighted) and the splice metric rank box plot^31^ (mis-splicing outlier in R072 highlighted). The AE and AS events resulted from a *de novo* hemizygous c.970G>A predicted missense variant, previously classified as a VUS^20^. RNA-seq sashimi plot shows skipping of exons 6 and 7 as well as retention of intron 7 as consequences of c.970G>A in R072 (blue), not observed in representative control fibroblasts (gray). The location of the *MBTPS2* variant is indicated by a triangle. In the schematic representation of the mis-splicing event, IGV^24^ screenshots of visualized WGS and RNA-seq data depict the read pileups at the mutant chromosomal position. (**D**) RNA-seq Manhattan plot representation of a 6q22.1-q22.31 microdeletion involving the dystonia-linked gene *NUS1* in patient R139. OUTRIDER^30^ identified significant AE (FCs: 0.44-0.59) of 6 genes in the affected chromosomal region 6q22.1-q22.31 (purple dots), including *NUS1* (FC: 0.51), supporting the presence of the pathogenic CNV in the patient. The X-axis represents chromosomes, and the Y-axis represents z-scores. AE, aberrant expression; AS, aberrant splicing; CNV, copy-number variant; DROP, Detection of RNA Outliers Pipeline; Ex, exon; FDR, false-discovery rate; FRASER, Find RAre Splicing Events in RNA-seq; hem, hemizygous; IGV, Integrative Genomics Viewer; LoF, loss-of-function; MAE, mono-allelic expression; MAF, minor allele frequency; n.t., not tested; OUTRIDER, Outlier in RNA-Seq Finder; RNA-seq, RNA sequencing; VUS, variant of uncertain significance; WES, whole-exome sequencing; WGS, whole-genome sequencing; wt, wild type.

### New diagnostic findings enabled by RNA-seq

Subsequently, variant (re)prioritization using our RNA-seq analysis pipeline was conducted on the “variant-negative” group based on prior WES/WGS sequencing results. RNA-seq aided in establishing new diagnoses in 9 index patients, providing a diagnostic uplift of 6.9% (9/131). Causative variants that had not been prioritized in genomic-data analysis alone were found through significant AS outliers (*n* = 3) or combined outliers (significant AS plus significant AE, *n* = 6) (**Tab.1**). No new diagnoses were made by MAE analysis. In 3 cases, RNA-seq directed the prioritization of extended splice-region variants (±3bp to ±10bp region) previously missed by WES/WGS^2, 3^. In patient R030 with dystonia and spastic ataxia, an AS event was found on *ACP33* (*SPG21*), and subsequent RNA-seq data visualization showed complete skipping of exon 4; WES/WGS-data reinspection revealed an underlying homozygous c.306+6T>A variant, allowing us to diagnose Mast syndrome (**Tab.1**; **Fig.5A**). The second case involved R054, a patient with dystonia, ataxia, and intellectual disability; RNA-seq identified AS of *ATG7*, associated with spinocerebellar ataxia-31, a neurodevelopmental disorder with multisystem involvement^25^. On re-review of R054’s WES/WGS data, we identified a heterozygous *ATG7* c.528+3A>G variant, associated with exon-8 skipping and intron retention, in combination with a c.1090G>A (p.Gly364Ser) missense substitution, which was initially not retained due to lacking additional *ATG7* coding variants. Parental testing demonstrated the bi-allelic status of the variants, and additional phenotyping revealed cerebellar hypoplasia and characteristic gingival hyperplasia^25^, confirming the diagnosis (**Tab.1**; **Fig.5B**). A third case, patient R089, presented with dystonia, developmental delay, and epilepsy and was found to harbor an AS event in *GLS*; with reanalysis-based detection of a corresponding homozygous c.1713-9C>G variant causing exon-16 extension as well as skipping of exons 16 and 17, we diagnosed *GLS*-associated developmental and epileptic encephalopathy-71 (**Tab.1**; **Fig.5C**). Our pipeline also helped to discover deep(er) intronic variants (> ±10bp from exon-intron boundaries), diagnosing another 6 cases that could not have been solved without RNA-seq information. Firstly, a heterozygous c.144-24A>G variant occurring in *trans* with a c.487G>T (p.Asp163Tyr) missense variant in *SHQ1* was prioritized from trio-WGS after AS- and AE-outlier analyses in patient R082. Both changes induced splicing defects (c.144-24A>G: exon-2 extension; c.487G>T: exon-5 skipping), resulting in a significant decrease in *SHQ1* expression (FC: 0.51); R082 presented with dystonia, chorea, and intellectual impairment, consistent with the diagnosis of *SHQ1*-related neurodevelopmental disorder (**Tab.1**; **Fig.6A**). Secondly, patient R140, who manifested developmental delay, cerebellar atrophy, and mixed movement abnormalities with stereotypies and dystonia, had AS and AE (FC: 0.23) hits for *SNX14*; WGS-data reevaluation indicated that the identified *SNX14* mis-splicing and underexpression resulted from a homozygous c.1108+28A>G variant, which led to extension of exon 12; available biochemical-testing results (elevated urine glycosaminoglycans) complemented the diagnosis of spinocerebellar ataxia-20 (**Tab.1**; **Fig.6B**). Thirdly, we clarified the disease cause in patient R158 from a consanguineous family with a similarly affected sibling with dystonia, cerebellar signs, and developmental delay; in the sibling, WES had previously prioritized a homozygous missense variant in *GRID2* (associated with spinocerebellar ataxia-18), regarded as likely causative for the observed phenotype^2^. Since the *GRID2* variant was not found in homozygosity upon follow-up targeted testing in R158, a different genetic etiology was considered: through RNA-seq we found AE of *AGTPBP1* (FC: 0.20), in combination with an AS hit in the same gene; reprioritization of quartet-WGS variants uncovered a homozygous deep intronic *AGTPBP1* c.1303-4238C>G alteration shared by both siblings, which induced the inclusion of a cryptic exon; *AGTPBP1*-related childhood-onset neurodegeneration with cerebellar atrophy was thus diagnosed (**Tab.1**; **Fig.7**). Our RNA-seq analysis pipeline further contributed, in three additional index patients, to the characterization of causative intronic variants with disruptive effects on expression and splicing in *ATM*, *SPG11*, and *UFC1*, which have been described in detail in a multi-omics companion manuscript^3^; for a summary, see **Tab.1**.

**Figure 5.**
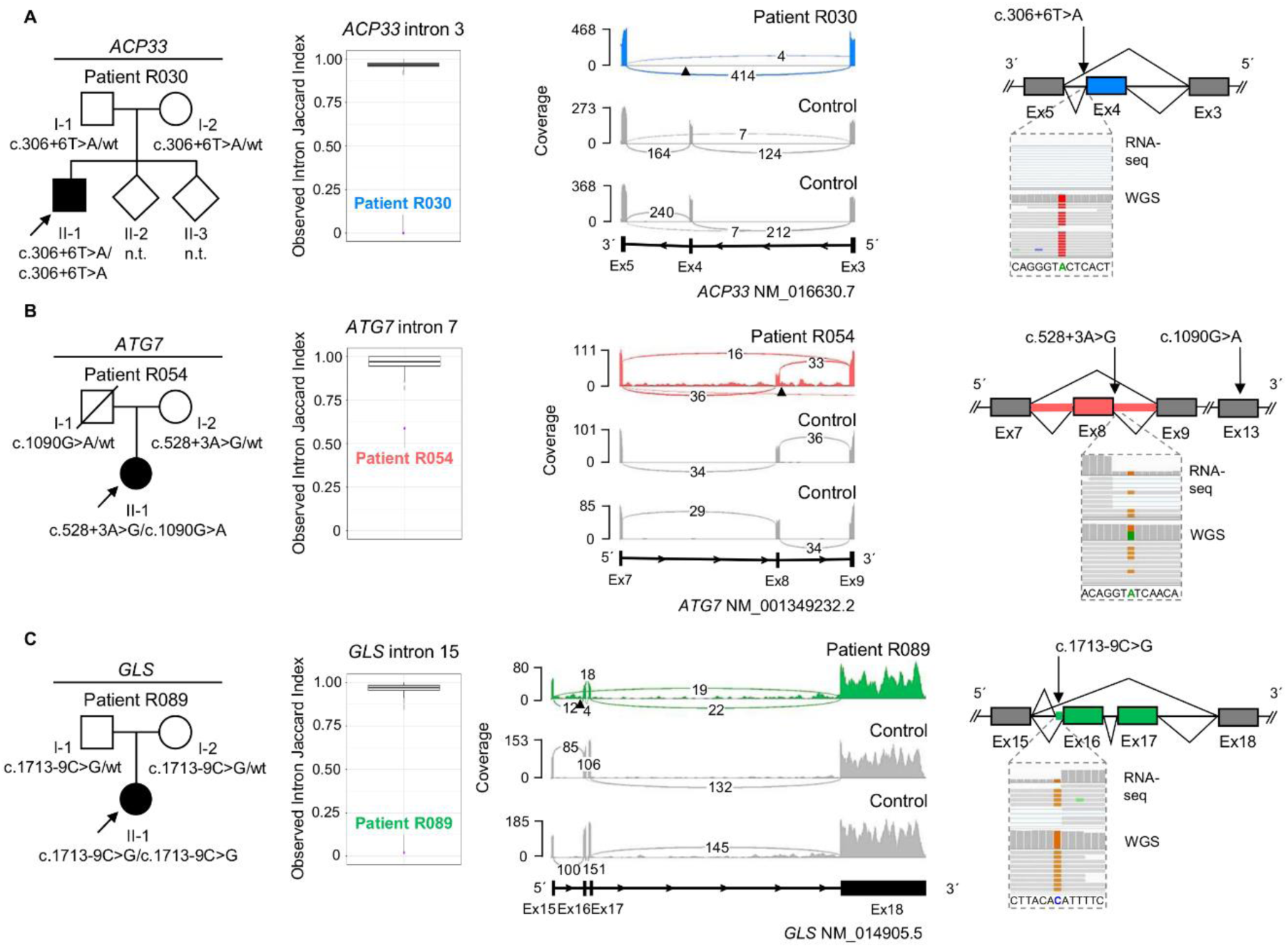
AS event-directed identification of causative extended splice-region variants. Pedigrees of patients R030 (**A**), R054 (**B**), and R089 (**C**) with suspected monogenic dystonic syndromes in whom WES/WGS alone did not provide a diagnosis^2, 3^. RNA-seq data-based significant splicing-outlier detection by FRASER 2.0^31^ guided the discovery of homozygous or compound heterozygous extended splice-region variants that were missed in the initial genomic analyses. Splice metric rank box plots^31^ highlight AS events for *ACP33* (*SPG21*, R030, blue), *ATG7* (R054, red), and *GLS* (R089, green). In the RNA-seq sashimi plots showing data from the patients and representative control fibroblasts (gray), the complete skipping of *ACP33* (*SPG21*) exon 4 as a consequence of the homozygous c.306+6T>A variant is in blue, the skipping of *ATG7* exon 8 as well as the retention of introns 7 and 8 as consequences of the heterozygous c.528+3A>G variant (found in *trans* with c.1090G>A) are in red, and the extension of *GLS* exon 16 as well as the skipping of exons 16 plus 17 as consequences of the homozygous c.1713-9C>G variant are in green. Variant locations are indicated by triangles. The mis-splicing events are also shown in the schematic representations, along with IGV^24^ screenshots of visualized WGS and RNA-seq data depicting the read pileups at the mutant chromosomal positions. AS, aberrant splicing; Ex, exon; FRASER, Find RAre Splicing Events in RNA-seq; IGV, Integrative Genomics Viewer; n.t., not tested; RNA-seq, RNA sequencing; WES, whole-exome sequencing; WGS, whole-genome sequencing; wt, wild type.

**Figure 6.**
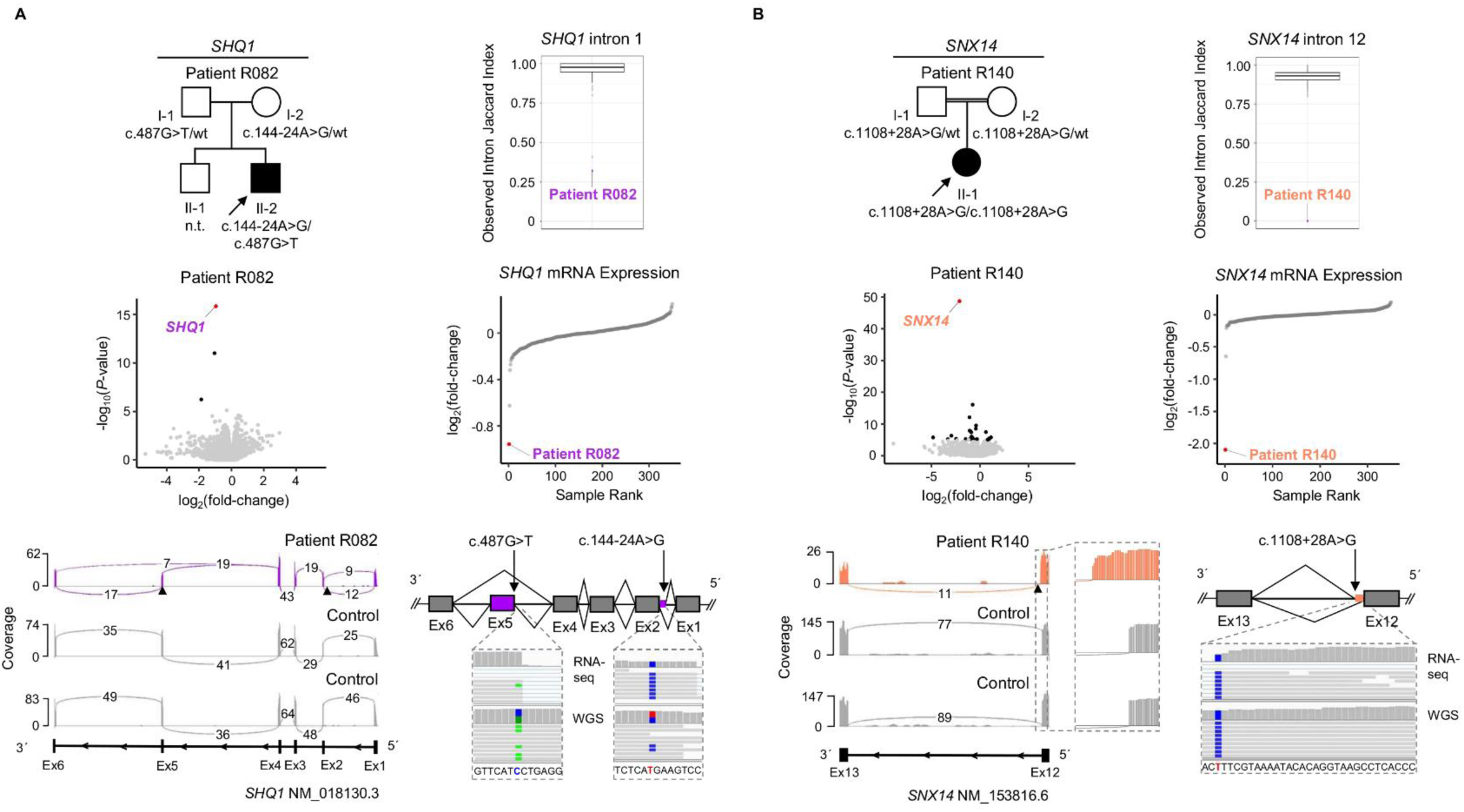
Examples of deep(er) intronic variant-related diagnoses enabled by automated RNA expression and splicing outlier analyses. Pedigrees of patients R082 (**A**) and R140 (**B**) with suspected monogenic dystonic syndromes in whom WES/WGS alone did not provide a diagnosis^2, 3^. RNA-seq data-based significant expression- and splicing-outlier detection by OUTRIDER^30^ and FRASER 2.0^31^ guided the discovery of compound heterozygous or homozygous deep(er) intronic variants that were not prioritized in the initial genomic analyses. Splice metric rank box plots^31^ highlight AS events, and RNA expression volcano and sample rank plots highlight AE events for *SHQ1* (R082, purple) and *SNX14* (R140, orange). In the RNA-seq sashimi plots showing data from the patients and representative control fibroblasts (gray), the extension of *SHQ1* exon 2 as a consequence of the heterozygous c.144-24A>G variant (found in *trans* with c.487G>T causing exon-5 skipping) is in purple, and the extension of *SNX14* exon 12 as a consequence of the homozygous c.1108+28A>G variant is in orange. A close-up of the *SNX14* exon-12 extension is provided. The mis-splicing events are also shown in the schematic representations, along with IGV^24^ screenshots of visualized WGS and RNA-seq data depicting the read pileups at the mutant chromosomal positions. AE, aberrant expression; AS, aberrant splicing; Ex, exon; FRASER, Find RAre Splicing Events in RNA-seq; IGV, Integrative Genomics Viewer; n.t., not tested; OUTRIDER, Outlier in RNA-Seq Finder; RNA-seq, RNA sequencing; WES, whole-exome sequencing; WGS, whole-genome sequencing; wt, wild type.

**Figure 7.**
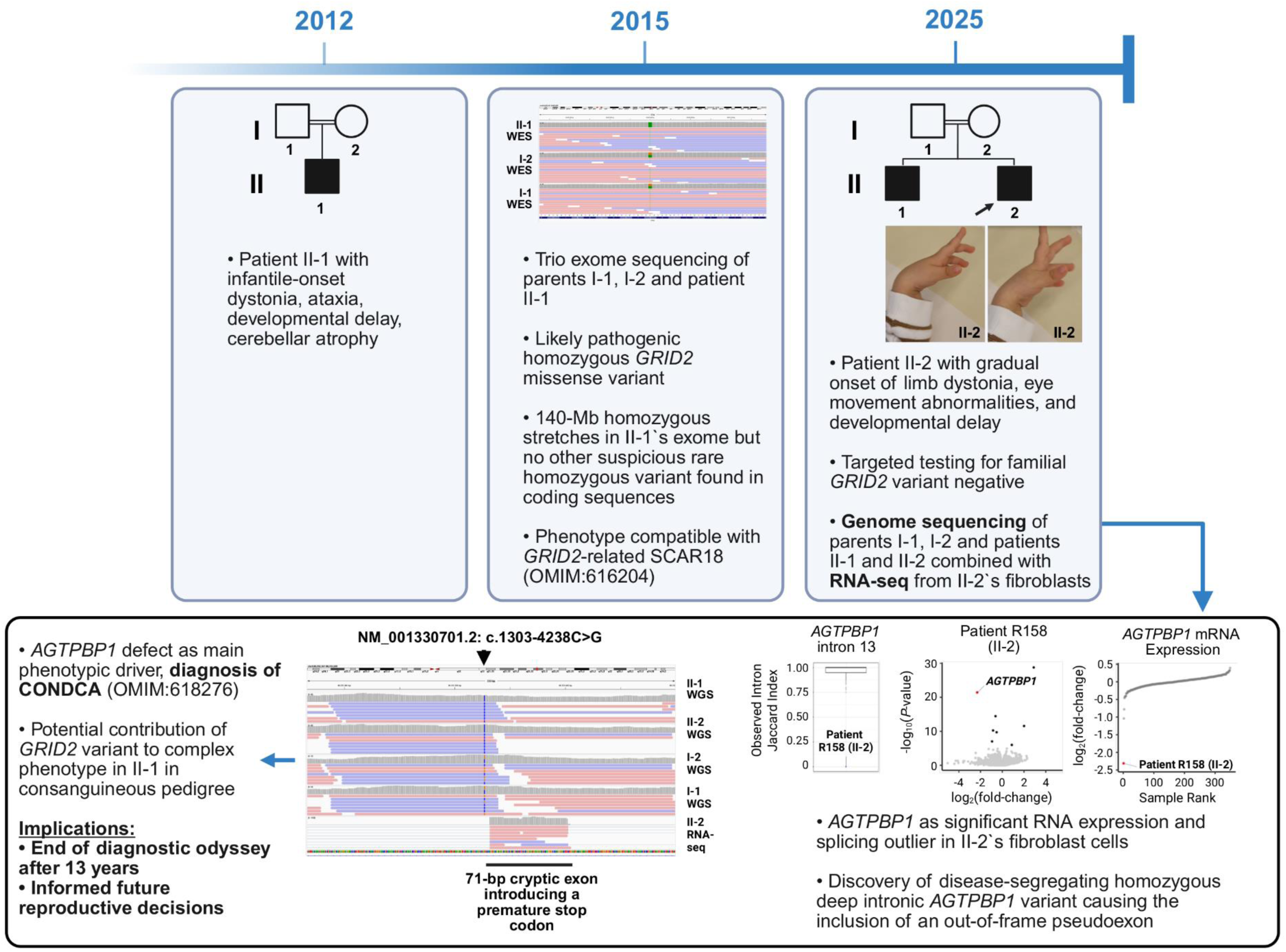
Example of the clinical value of RNA-seq: bringing an end to the diagnostic odyssey after 13 years. A timeline of the clinical course and applied molecular diagnostics in two similarly affected siblings, including patient R158 in this study (II-2 in the family pedigree, arrow; see II-2’s limb dystonia in the clinical photographs). An originally WES-prioritized homozygous *GRID2* missense variant considered explanatory for the phenotype of the older brother was not identified in R158, ruling out its role as the true (sole) disease-causing variant in the family. RNA-seq and WGS reanalysis were instrumental in uncovering the shared monogenic etiology in the siblings: significant AE and AS events^17^ were found for *AGTPBP1* in R158 (II-2)’ fibroblasts, resulting from the activation of a pseudoexon through a homozygous deep intronic c.1303-4238C>G variant (highlighted by a triangle in the IGV^24^ screenshot, bottom box). Corresponding splice metric rank box plot^31^ as well as RNA expression volcano and sample rank plots are shown, illustrating the *AGTPBP1* outlier events. The results influenced reproductive decision-making after a 13-year diagnostic journey. AE, aberrant expression; AS, aberrant splicing; CONDCA, neurodegeneration, childhood-onset, with cerebellar atrophy; IGV, Integrative Genomics Viewer; RNA-seq, RNA sequencing; WES, whole-exome sequencing; WGS, whole-genome sequencing.

### Incorporation of AbExp scores

Investigating the consequences of dystonia-associated variants on gene expression in the relevant brain regions promotes a better understanding of molecular mechanisms^1^. Our study scope only included tissue obtainable in a clinical setting, so we applied the machine-learning model AbExp^18^, which predicts aberrant underexpression in non-accessible tissues, to causative variants that led to significantly reduced RNA abundance of affected genes in fibroblasts. This approach estimated that most of the variants for which scores could be obtained from AbExp^18^ (22/25, 88.0%) would operate by loss of expression in different brain regions, including those relevant to dystonia, although the obtained *z*-scores did not reach the low-/high-confidence underexpression threshold in 62.6% of all predictions (**Suppl.Tab.2**). Integrating our present fibroblast RNA-seq AE results improved the variant-level predictions, such that combined analysis predicted relevant underexpression in different brain regions for all variants (71.5% high-confidence underexpression outliers, 28.5% low-confidence underexpression ouliers; **Suppl.Tab.2**), further supporting the use of fibroblast-based RNA-seq as an effective strategy for diagnostic evaluation of dystonia.

## Discussion

Our study provides a systematic evaluation of the diagnostic potential of fibroblast RNA-seq for a heterogeneous group of dystonic diseases, using a streamlined analysis pipeline. Establishing multiomics pipelines is essential, as standard WES/WGS approaches often fail to provide a diagnosis for many individuals affected by dystonia^2, 3^. Our present framework indicates that (1) fibroblasts are suitable for transcriptomic assessment of a broad spectrum of associated genes and (2) advanced bioinformatic tools^17, 30, 31^ yield a reviewable set of candidate outliers. While the present exploration focused on a unique set of patients with (ultra)rare dystonic syndromes and diagnostically intractable conditions, recruited to add comprehensive molecular-characterization analyses in the context of suspected monogenic causation, we expect that RNA-seq is extendable to the wider dystonia population for unraveling pathogenic variant consequences and genetic etiologies—especially difficult-to-interpret non-coding alterations. Motivated by applications in other rare-disease cohorts^12–16^, we evaluated the detection of a dystonia-associated gene set in fibroblast RNA-seq, finding a substantial proportion of genes linked to dystonia (>80% of OMIM dystonia genes^25^). Beyond the utility of fibroblast material for discovering mutational downstream effects in a wide spectrum of genetic subtypes, it promotes evaluation of genes organized into disease pathways^29^, helping resolve the role of variants in yet-to-be-discovered dystonia-causal genes impacting converging processes. Although more invasive, fibroblasts expressed a higher percentage of genes relevant to dystonia than blood. Evidently, skin biopsies may represent the recommended source for RNA-seq-based diagnostics in dystonia. Integrating the widely used DROP workflow^17^, each investigated patient sample had a maximum of 3 significant outlier events in OMIM dystonia genes^25^. This low number of hits made manual review effective, demonstrating that the profiling of expression and splicing outliers by algorithms such as OUTRIDER^30^ and FRASER 2.0^31^ could become a valuable component in the determination of variant mechanisms and the search for elusive pathogenic variants in dystonia. We assessed the capacity of our RNA-seq analysis pipeline to resolve RNA defects associated with pre-identified variants. We replicated previous observations^11, 14^ that many LoF variants (bi- and mono-allelic, 10/15, 66.7%) caused significant AE detectable by OUTRIDER, suggesting that RNA-seq and outlier analysis can successfully be used for uncovering dystonia-relevant underexpression and pathogenic mechanisms such as haploinsufficiency (e.g., for *VPS16*-related dystonia, 3/3 cases; **Fig.4A/B**; **Suppl.Tab.1**). While our stringent significance cutoff did not retain AE events for 4 cases with heterozygous LoF variants, additional sample rank-based evaluation^3, 32^ constituted a useful complementary strategy. Several other mutation types causally involved in dystonic syndromes, including missense variants, CNVs, repeat expansions, and synonymous changes, variably impacted measurable transcriptional outcomes according to our pipeline cutoffs, with detection of AE/AS events in 5/21 samples (23.8%; **Fig.4A**; **Suppl.Tab.1**). Surprising discoveries emerged from the use of RNA-seq, DROP, and visual inspection in cases with splice- and/or expression-disrupting missense (patient R072) and synonymous (R047) variants, expanding the genotype-phenotype spectra for the less-characterized genes *MBTPS*2^43^ and *HCN2*^44^. To prioritize variants evading prior identification by WES/WGS alone, we tested samples with hard-to-resolve dystonic conditions. This outlier-aided approach^14, 23^ with subsequent genomic reanalysis facilitated additional diagnoses in 6.9% of cases (9/131). Overall, the greatest success occurred in identifying WES/WGS-unprioritized causative variants in early-onset cases affected by autosomal recessive disorders in which dystonia was combined with other features. Positive findings were driven by AS and AE outliers resulting from “missed” variations in non-coding regions, promoting investigation into these regions through RNA-seq-supported strategies to better understand the elusive genetic basis of rare dystonias. New RNA-seq-based diagnoses had clinical implications. For example, in patient R158’s family (**Fig.7**), the discovery of a causative deep-intronic homozygous *AGTPBP1* variant informed future reproductive decisions. Lastly, the machine learning tool, AbExp^18^, predicted that the variants associated with AE in skin would decrease the expression of affected genes in different dystonia-relevant brain regions. Subsequently, more comprehensive incorporation of such computer-aided modeling will increase the sensitivity for prioritization of variants that affect RNA integrity in neurological disorders, such as dystonia, in the future, especially given limited easy access to neuronal tissue^45^. Limitations of this study included a relatively small cohort size and a restricted set of dystonia-associated variants and VUSs available for benchmarking analysis. Although our pipeline covered many genes, not all relevant genes could be tested due to non-expression in fibroblasts (∼20%). Moreover, we cannot exclude that potentially more sensitive methods for the discovery of RNA defects would outperform our present results, selecting DROP for its current performance, ease of use, and interpretability of results. Our data ultimately encourages the adaptation of RNA-seq for deepening our understanding of mutational mechanisms and improving molecular diagnostics in dystonic diseases.

## Data Availability

All data produced in the present study are available upon reasonable request to the authors.

## Acknowledgements

This work was supported by a “*Schlüsselprojekt*” grant from the Else Kröner-Fresenius-Stiftung (2022_EKSE.185). This research was also supported by funding from the EJP RD (EJP RD Joint Transnational Call 2022) and the German Federal Ministry of Education and Research (BMBF, Bonn, Germany), awarded to the project PreDYT (PREdictive biomarkers in DYsTonia, 01GM2302). The work was further supported by the DFG Research Infrastructure NGS_CC (project #458949627) as part of the Next Generation Sequencing Competence Network (project 423957469); NGS analyses were carried out at the production site WGGC-Bonn; J.W. and M.Z. received the DFG grants WI 1820/14-1 and ZE 1213/2-1 (#458949627) as part of the DFG Sequencing call Sequencing Costs in Projects. In addition, this study (M.Z.) has received funding from the Federal Ministry of Education and Research (BMBF) and the Free State of Bavaria under the Excellence Strategy of the Federal Government and the Länder, as well as by the Technical University of Munich - Institute for Advanced Study. Part of this research was made possible through access to the data (Technical University of Munich, Munich, Germany) generated by the Bavarian Genomes Network. H.P. acknowledges grant support from the German Federal Ministry of Education and Research (BMBF, Bonn, Germany) awarded to the German Network for Mitochondrial Disorders (mitoNET, 01GM1906A). H.P. also received grant support from the EJP RD and the BMBF (Bonn, Germany), awarded to the project GENOMIT (01GM2301 and 01GM2404A). M.S. received support from the EU Renewal and Resilience Plan “Large projects for excellent researchers” under grant No. 09I03-03-V03-0000. This study was supported by the Czech Ministry of Health – grant AZV NW24-04-00067 and the project National Institute for Neurological Research (Programme EXCELES, ID Project No. LX22NPO5107) – Funded by the European Union – Next Generation EU. The IT infrastructure at the Chair of CMM (TUM) was co-funded by the Deutsche Forschungsgemeinschaft (DFG, German Research Foundation) – Project-IDs 461264291 and 553375143. ERDERA has received funding from the European Union’s Horizon Europe research and innovation programme under grant agreement N°101156595 (V.A.Y.). Views and opinions expressed are those of the author(s) only and do not necessarily reflect those of the European Union or any other granting authority, who cannot be held responsible for them. This work was supported by the EXIST programme of the German Federal Ministry for Economic Affairs and Climate Action (BMWK) and co-funded by the European Union (03EGTBY028 to V.A.Y., C.M., and F.B.). Several authors of this paper are members of the European Reference Network for Rare Neurological Diseases (Project ID 739510). The authors would like to thank the patients and their families for their participation in this study. We thank Katharina Eyring, Katharina Mayerhanser, Nicola Plathner, and Silke Slanz for excellent technical assistance. M.Z. is a member of the Medical and Scientific Advisory Council of the Dystonia Medical Research Foundation (DMRF) and a member of the Governance Council of the International Cerebral Palsy Genomics Consortium (ICPGC). BioRender was used to create (parts of) figures 1 and 7 (https://www.biorender.com).

## Author contributions

A.S., I.D., R.J., J.W., E.G., J.G., M.S., and M.Z. contributed to the conception and design of the study. A.S., I.D., T.B., V.A.Y., F.H., E.I., B.A., S.B., D.B., S.B., F.B., M.B., K.D., F.D., P.H., D.H., P.H., A.-K. J., M.K., R.K., A.K., M.K., L.K., K.K., O.M., M.M-B., C.M., A.O., T.R., A.S., U.S., A. M. S., K.V., M.W., H.P., S.B., J.N., R.J., J.W., E.G., J.G., M.S., and M.Z. contributed to the acquisition and/or analysis of the data. A.S., I.D., and M.Z. contributed to drafting the text and/or preparing the figures.

