## Supplemental Table 1 and 2 for "Fibroblast transcriptomics in molecular diagnostics of a comprehensive dystonia cohort"

### Table of contents

#### Supplementary Online Table


| | |
|---|---|
| Supplementary Online Table 1 | Summary of pre-identified variants investigated by RNA-seq in this study |
| Supplementary Online Table 2 | Summary of AbExp scores for dystonia-associated variants causing underexpression in skin fibroblasts |

### Supplementary Online Table

**Supplementary Online Table 1** Summary of pre-identified variants[1] investigated by RNA-seq in this study

| Index patient[7]/ genomic sequencing | Phenotype | Gene/ associated disorder (OMIM) | Gene expressed in fibroblasts | Variant(s) (zygosity, clinical significance category: LP/P versus VUS) | RNA phenotype | Tool(s) that identified significant RNA defect | RNA phenotype supporting clinical evaluation of the variant(s) |
|---|---|---|---|---|---|---|---|
| **Bi-allelic LoF variants** | | | | | | | |
| R028/ WES+WGS | dystonia, cognitive decline | *POLR3A*/ leukodystrophy, hypomyelinating, 7, with or without oligodontia and/or hypogonadotropic hypogonadism (607694) | yes | NM_007055.4: c.1771-6C>G, p.? (hom, LP/P) | *POLR3A* significant underexpression (FC: 0.57)[3], exon-14 skipping | OUTRIDER | yes: support of LoF effect, support of splice defect |
| R023/ WES | dystonia, DD | *ZNF142*/ neurodevelopmental disorder with impaired speech and hyperkinetic movements (618425) | yes | NM_001105537.4: c.3175C>T, p.Arg1059* (hom, LP/P) | *ZNF142* significant underexpression (FC: 0.67) | OUTRIDER | yes: support of LoF effect |
| **Mono-allelic LoF variants** | | | | | | | |
| R029/ WES | dystonia, chorea | *ADCY5*/ dyskinesia with orofacial involvement, autosomal dominant (606703) | no | NM_183357.2: c.2088+1G>A, p.? (het, LP/P) | N/A | N/A | no |
| R020/ WES | dystonia, psychiatric features | *ANK2*/ *ANK2*-related neurodevelopmental disorder (N/A) | yes | NM_001148.6: c.3804dup, p.Thr1269Hisfs*19 (het, LP/P) | N/A[2] | N/A | (no)[2] |
| R161/ WES+WGS | dystonia, myoclonus, spasticity, DD | *ATP5F1A*/ mitochondrial complex V (ATP synthase) deficiency, nuclear type 4A (620358) | yes | NM_004046.6: c.1404del, p.Glu469Serfs*3 (het, LP/P) | *ATP5F1A* significant underexpression (FC: 0.63)[3] | OUTRIDER, MAE | yes: support of LoF effect/ haploinsufficiency mechanism |

| | | | | | | | |
|---|---|---|---|---|---|---|---|
| R016/ WES+WGS | dystonia, DD | *ATP5F1B*/ *ATP5F1B*-related dystonia (N/A) | yes | NM_001686.4: c.1074+1G>T, p.? (het, LP/P) | *ATP5F1B* significant underexpression (FC: 0.69)[3], exon-7 skipping, skipping of exons 6 and 7 | OUTRIDER | yes: support of LoF effect/ haploinsufficiency mechanism, support of splice defect |
| R050/ WES | dystonia | *CHD3*/ Snijders Blok-Campeau syndrome (618205) | yes | NM_001005273.3: c.793+1G>A, p.? (het, LP/P) | *CHD3* significant underexpression (FC: 0.65), exon-5 skipping | OUTRIDER, FRASER 2.0 | yes: support of LoF effect/ haploinsufficiency mechanism, support of splice defect |
| R051/ WES | dystonia | *EIF4A2*/ neurodevelopmental disorder with hypotonia and speech delay, with or without seizures (620455) | yes | NM_001967.4: c.896_897del, p.Thr299Serfs*7 (het, LP/P) | *EIF4A2* significant underexpression (FC: 0.74) | OUTRIDER, MAE | yes: support of LoF effect/ haploinsufficiency mechanism |
| R095/ WES+WGS | dystonia, ID, epilepsy | *IRF2BPL*/ neurodevelopmental disorder with regression, abnormal movements, loss of speech, and seizures (618088) | yes | NM_024496.4: c.189_211dup, p.Gly71Alafs*89 (het, LP/P) | N/A[4] | N/A | no |
| R154/ WES | dystonia, DD, microcephaly | *KMT2B*/ dystonia 28, childhood-onset (617284) | yes | NM_014727.2: c.610C>T, p.Gln204* (het, LP/P) | N/A[5] | N/A | no |
| R146/ WES+WGS | dystonia, ataxia, spasticity, ID, epilepsy | *MECP2*/ Rett syndrome (312750) | yes | NM_004992.4: c.1146_1193delinsC, p.Leu383Profs*6 (het, LP/P) | N/A[6] | N/A | no |
| R083/ WES+WGS | dystonia, spasticity | *PTPN1*/ *PTPN1*-associated interferonopathy (N/A) | yes | NM_002827.4: c.505C>T, p.Arg169* (het, LP/P) | *PTPN1* significant underexpression (FC: 0.65) | OUTRIDER, MAE | yes: support of LoF effect/ haploinsufficiency mechanism |
| R044/ WES | dystonia, psychiatric features | *VPS16*/ dystonia 30 (619291) | yes | NM_022575.4: c.559C>T, p.Arg187* (het, LP/P) | *VPS16* significant underexpression (FC: 0.58) | OUTRIDER, MAE | yes: support of LoF effect/ haploinsufficiency mechanism |
| R104/ WES | dystonia | *VPS16*/ dystonia 30 (619291) | yes | NM_022575.4: c.1988_1989insG, p.Asn663Lysfs*2 (het, LP/P) | *VPS16* significant underexpression (FC: 0.59) | OUTRIDER, MAE | yes: support of LoF effect/ haploinsufficiency mechanism |
| R137/ WES | dystonia, psychiatric features | *VPS16*/ dystonia 30 (619291) | yes | NM_022575.4: c.559C>T, p.Arg187* (het, LP/P) | *VPS16* significant underexpression (FC: 0.59) | OUTRIDER | yes: support of LoF effect/ haploinsufficiency mechanism |

| | | | | | | | |
|---|---|---|---|---|---|---|---|
| **Bi-allelic missense variants** | | | | | | | |
| R021/ WES | dystonia, ataxia, DD, microcephaly | *NUP54*/ dystonia 37, early-onset, with striatal lesions (620427) | yes | NM_017426.4: c.1073T>G, p.Ile358Ser (hom, LP/P) | N/A | N/A | no |
| R114/ WES | dystonia, myoclonus, ataxia, renal abnormality | *SLC30A9*/ Birk-Landau-Perez syndrome (617595) | yes | NM_006345.4: c.896C>T, p.Pro299Leu + c.1484A>G, p.Asp495Gly (comp het, VUS + VUS) | N/A | N/A | no |
| **Mono-allelic/hemizygous missense variants** | | | | | | | |
| R031/ WES+WGS | dystonia, ataxia | *ATP2B2*/ *ATP2B2*-related neurodevelopmental disorder (N/A) | no | NM_001001331.4: c.3028G>A, p.Glu1010Lys (het, LP/P) | N/A | N/A | no |
| R141/ WES | dystonia | *ATP5MC3*/ dystonia, early-onset, and/or spastic paraplegia (619681) | yes | NM_001689.5: c.318C>G, p.Asn106Lys (het, LP/P) | N/A | N/A | no |
| R132/ WES+WGS | dystonia, myoclonus, DD, ID, epilepsy | *DNM1L*/ encephalopathy, lethal, due to defective mitochondrial peroxisomal fission 1 (614388) | yes | NM_012062.5: c.176C>T, p.Thr59Ile (het, LP/P) | N/A | N/A | no |
| R052/ WES | dystonia, ID, epilepsy, microcephaly | *MATR3*/ *MATR3*-related early-onset neurodegeneration (N/A) | yes | NM_018834.6: c.1306G>A, p.Glu436Lys (het, LP/P) | N/A | N/A | no |
| R072/ WES+WGS | dystonia, DD, epilepsy | *MBTPS2*/ IFAP syndrome with or without BRESHECK syndrome (308205) | yes | NM_015884.4: c.970G>A, p.Ala324Thr (hem, VUS) | *MBTPS2* significant underexpression (FC: 0.65), skipping of exons 6 and 7, partial retention of intron 7 | OUTRIDER, FRASER 2.0 | yes: demonstration of LoF effect, demonstration of splice defect, variant re-classified based on RNA defect (VUS > LP/P) |
| R130/ WES+WGS | dystonia, myoclonus | *SGCE*/ dystonia-11, myoclonic (159900) | yes | NM_003919.3: c.742T>A, p.Cys248Ser (het, LP/P) | N/A | N/A | no |
| R085/ WES+WGS | dystonia, ataxia, myoclonus, DD, ID | *SLC16A2*/ Allan-Herndon-Dudley syndrome (300523) | yes | NM_006517.5: c.1025T>C, p.Leu342Pro (het, LP/P) | N/A | N/A | no |

| | | | | | | | |
|---|---|---|---|---|---|---|---|
| **CNVs** | | | | | | | |
| R139/ WES+WGS | dystonia, myoclonus, DD | *NUS1*/ intellectual developmental disorder, autosomal dominant 55, with seizures (617831) | yes | chr6:117571001-122124500, del chr6q22.1-q22.31, multiple genes including *NUS1* (het, LP/P) | significant underexpression of *NUS1* (FC: 0.51) and 5 other genes in chromosomal region chr6q22.1-q22.31 | OUTRIDER | yes: support for the presence of deletion in region chr6q22.1-q22.31, support of LoF effect/ haploinsufficiency mechanism (*NUS1*) |
| R135/ WES+WGS | dystonia, ataxia, spasticity | *SPG7*/ spastic paraplegia 7, autosomal recessive (607259) | yes | chr16:89596808-89597521, del exon 7 (NM_003119.4) (hom, LP/P) | *SPG7 e*xon-7 skipping | FRASER 2.0 | yes: support for the presence of single-exon deletion |
| **Repeat expansions** | | | | | | | |
| R057/ WES+WGS | dystonia, ataxia | *ATXN8OS* (*SCA8*)/ spinocerebellar ataxia 8 (608768) | no | chr13:70713515[124] (NR_185841.1), CI: 95-174 CTG units (het, pathological range) | N/A | N/A | no |
| R010/ WES+WGS | dystonia, ataxia, myoclonus, epilepsy | *CSTB*/ epilepsy, progressive myoclonic 1A, Unverricht and Lundborg (254800) | yes | chr21:45196349[38] (NM_000100.4), CI: 27-51 CCCCGCCCCGCG units (allele 1) and 22-42 CCCCGCCCCGCG units (allele 2) (hom, pathological range) | *CSTB* significant underexpression (FC: 0.31)[3] | OUTRIDER | yes: support for presence of repeat expansion, support of LoF effect |
| R100/ WES+WGS | dystonia | *DIP2B*/ *DIP2B*-related movement disorder (N/A) | yes | chr12:50898784[80] (NM_173602.3), CI: 68-137 GGC units (het, pathological range) | N/A | N/A | no |
| R152/ WES+WGS | dystonia | *FMR1*/ fragile-X-associated tremor/ataxia syndrome (300623) | yes | chrX:146993568[92] (NM_002024.6), CI: 79-159 CGG units (het, pathological range) | N/A | N/A | no |
| R034/ WES+WGS | dystonia, ataxia, DD, ID | *GLS*/ global developmental delay, progressive ataxia, and elevated glutamine (618412) | yes | chr2:191745598[138] (NM_014905.5), CI: 115-195 GCA units + c.1197+2T>C, p.? (comp | *GLS* significant underexpression (FC: 0.59), exon-10 extension (c.1197+2T>C) | OUTRIDER, FRASER 2.0 | yes: support for presence of repeat expansion, support of splice defect (c.1197+2T>C), support of LoF effect |

| | | | | | | | |
|---|---|---|---|---|---|---|---|
| | | | | het, pathological range + LP/P) | | | |
| R151/ WES+WGS | dystonia, chorea | *HTT*/ Huntington disease (143100) | yes | chr4:3076603[40] (NM_001388492.1), 40 CAG units (het, pathological range) | N/A | N/A | no |
| R101/ WES+WGS | dystonia, chorea, cognitive decline, muscle wasting | *PABPN1*/ oculopharyngeal muscular dystrophy-1 (164300) | yes | chr14:23790681[7] (NM_004643.4), 4 GCG and 3 GCA units (het, pathological range) | N/A | N/A | no |
| **Synonymous variants (in *trans* with other variant types)** | | | | | | | |
| R047/ WES+WGS | dystonia, epilepsy, DD | *HCN2*/ *HCN2*-related neurodevelopmental disorder, autosomal recessive (N/A) | yes | NM_001194.4: c.1560C>T, p.Gly520= + del exon 6 (comp het, VUS + LP/P) | IGV-based manual inspection: cryptic splice donor created by c.1560C>T with exon-5 truncation (only visible in 3 reads due to NMD), lowest *HCN2* expression in sample-rank analysis | N/A | yes: support of LoF effect (sample-rank analysis), demonstration of splice defect (IGV), synonymous variant re-classified based on RNA defect (VUS > LP/P) |
| R149/ WES+WGS | dystonia, DD, metabolic decompensation | *SARS2*/ hyperuricemia, pulmonary hypertension, renal failure, and alkalosis (613845) | yes | NM_017827.4: c.446T>C, p.Leu149Pro + c.627C>T, p.Gly209= (comp het, VUS + VUS) | N/A | N/A | no |
| R093/ WES+WGS | dystonia, ataxia, ID | *TARS2*/ combined oxidative phosphorylation deficiency 21 (615918) | yes | NM_025150.5: c.774G>T, p.Ser258= + c.1099C>T, p.His367Tyr (comp het, VUS + VUS) | N/A | N/A | no |

[1]27 of 36 cases (75.0%) with pre-identified variants were previously published by us: PMIDs: 39937650, 31036918, 40276935, 37485550, 39986310, 32808683, 33998058, 36333996, 34173818, 34954817, 40590478, 37675773.
[2]Lowest *ANK2* expression compared to all other samples that expressed the gene in sample-rank analysis (sample rank: 1/349, FC: 0.51, FDR>0.05).
[3]FC calculated based on new RNA-seq dataset compared to PMIDs: 40276935 and 39937650.
[4]*IRF2BPL* is a single-exon gene and NMD is not expected.

[5]*KMT2B* not significantly underexpressed (sample rank: 25/349, FC: 0.94, FDR>0.05), index patient with milder (atypical) phenotype.
[6]*MECP2* variant located in the last exon with possible NMD escape (sample rank: 60/349, FC: 0.97, FDR>0.05).
[7]**The patient IDs were not known to anyone outside the research group.**

Abbreviations: CI, confidence interval (ExpansionHunter); CNV, copy number variant; comp het, compound heterozygous; DD, developmental delay; del, deletion; F, female; FC, fold change; FDR, false-discovery rate; FRASER, Find RAre Splicing Events in RNA-seq; hem, hemizygous; het, heterozygous; hom, homozygous; ID, intellectual disability; IGV, Integrative Genomics Viewer; LoF, loss-of-function; LP/P, likely pathogenic/pathogenic; MAE, mono-allelic expression; M, male; N/A, not applicable/not available; NMD, nonsense-mediated mRNA decay; OMIM, Online Mendelian Inheritance in Man; OUTRIDER, Outlier in RNA-Seq Finder; RNA-seq, RNA sequencing; VUS, variant of uncertain significance, WES, whole-exome sequencing; WGS, whole-genome sequencing; y, years,

**Supplementary Online Table 2** Summary of AbExp scores for dystonia-associated variants causing underexpression in skin fibroblasts

| Chromosome | Start | End | Ref | Alt | Gene | Tissue | Tissue Type | AbExp (z-score) | RNA AbExp (logistic regression) |
|---|---|---|---|---|---|---|---|---|---|
| chr1 | 161127085 | 161127098 | TGAGTTTGACATC | T | ENSG00000143222 | Brain - Caudate (basal ganglia) | Brain | -0.1525284 | 0.49196146 |
| chr1 | 161127085 | 161127098 | TGAGTTTGACATC | T | ENSG00000143222 | Brain - Cerebellar Hemisphere | Brain | -0.1525284 | 0.49196146 |
| chr1 | 161127085 | 161127098 | TGAGTTTGACATC | T | ENSG00000143222 | Brain - Cortex | Brain | -0.1525284 | 0.49196146 |
| chr1 | 161127085 | 161127098 | TGAGTTTGACATC | T | ENSG00000143222 | Brain - Frontal Cortex (BA9) | Brain | -0.1525284 | 0.49196146 |
| chr1 | 161127085 | 161127098 | TGAGTTTGACATC | T | ENSG00000143222 | Brain - Hippocampus | Brain | -0.1525284 | 0.49196146 |
| chr1 | 161127085 | 161127098 | TGAGTTTGACATC | T | ENSG00000143222 | Brain - Hypothalamus | Brain | -0.1525284 | 0.49196146 |
| chr1 | 161127085 | 161127098 | TGAGTTTGACATC | T | ENSG00000143222 | Brain - Nucleus accumbens (basal ganglia) | Brain | -0.1525284 | 0.49196146 |
| chr1 | 161127085 | 161127098 | TGAGTTTGACATC | T | ENSG00000143222 | Brain - Putamen (basal ganglia) | Brain | -0.1525284 | 0.49196146 |
| chr1 | 161127085 | 161127098 | TGAGTTTGACATC | T | ENSG00000143222 | Brain - Spinal cord (cervical c-1) | Brain | -0.1525284 | 0.49196146 |
| chr1 | 161127085 | 161127098 | TGAGTTTGACATC | T | ENSG00000143222 | Brain - Substantia nigra | Brain | -0.1512097 | 0.49194345 |
| chr1 | 161127123 | 161127124 | G | A | ENSG00000143222 | Brain - Caudate (basal ganglia) | Brain | -0.1285439 | 0.49163396 |
| chr1 | 161127123 | 161127124 | G | A | ENSG00000143222 | Brain - Cerebellar Hemisphere | Brain | -0.1396183 | 0.49178518 |
| chr1 | 161127123 | 161127124 | G | A | ENSG00000143222 | Brain - Cortex | Brain | -0.1285439 | 0.49163396 |
| chr1 | 161127123 | 161127124 | G | A | ENSG00000143222 | Brain - Frontal Cortex (BA9) | Brain | -0.1285439 | 0.49163396 |
| chr1 | 161127123 | 161127124 | G | A | ENSG00000143222 | Brain - Hippocampus | Brain | -0.1285439 | 0.49163396 |
| chr1 | 161127123 | 161127124 | G | A | ENSG00000143222 | Brain - Hypothalamus | Brain | -0.1285439 | 0.49163396 |
| chr1 | 161127123 | 161127124 | G | A | ENSG00000143222 | Brain - Nucleus accumbens (basal ganglia) | Brain | -0.1285439 | 0.49163396 |
| chr1 | 161127123 | 161127124 | G | A | ENSG00000143222 | Brain - Putamen (basal ganglia) | Brain | -0.1285439 | 0.49163396 |
| chr1 | 161127123 | 161127124 | G | A | ENSG00000143222 | Brain - Spinal cord (cervical c-1) | Brain | -0.1285439 | 0.49163396 |
| chr1 | 161127123 | 161127124 | G | A | ENSG00000143222 | Brain - Substantia nigra | Brain | -0.1272251 | 0.49161595 |
| chr2 | 191788710 | 191788711 | T | C | ENSG00000115419 | Brain - Caudate (basal ganglia) | Brain | -2.704433 | 0.24553125 |
| chr2 | 191788710 | 191788711 | T | C | ENSG00000115419 | Brain - Cerebellar Hemisphere | Brain | -2.8238558 | 0.2574249 |
| chr2 | 191788710 | 191788711 | T | C | ENSG00000115419 | Brain - Cortex | Brain | -3.3163621 | 0.31028616 |
| chr2 | 191788710 | 191788711 | T | C | ENSG00000115419 | Brain - Frontal Cortex (BA9) | Brain | -2.3989541 | 0.21682993 |
| chr2 | 191788710 | 191788711 | T | C | ENSG00000115419 | Brain - Hippocampus | Brain | -2.1689678 | 0.19687586 |

| | | | | | | | | | |
|---|---|---|---|---|---|---|---|---|---|
| chr2 | 191788710 | 191788711 | T | C | ENSG00000115419 | Brain - Hypothalamus | Brain | -2.2358091 | 0.20252838 |
| chr2 | 191788710 | 191788711 | T | C | ENSG00000115419 | Brain - Nucleus accumbens (basal ganglia) | Brain | -2.0701201 | 0.18873634 |
| chr2 | 191788710 | 191788711 | T | C | ENSG00000115419 | Brain - Putamen (basal ganglia) | Brain | -2.3203888 | 0.20985326 |
| chr2 | 191788710 | 191788711 | T | C | ENSG00000115419 | Brain - Spinal cord (cervical c-1) | Brain | -1.4335609 | 0.14245065 |
| chr2 | 191788710 | 191788711 | T | C | ENSG00000115419 | Brain - Substantia nigra | Brain | -1.8080277 | 0.16841104 |
| chr2 | 219508063 | 219508064 | G | A | ENSG00000115568 | Brain - Caudate (basal ganglia) | Brain | -3.29402 | 0.49736537 |
| chr2 | 219508063 | 219508064 | G | A | ENSG00000115568 | Brain - Cerebellar Hemisphere | Brain | -2.3077184 | 0.46547442 |
| chr2 | 219508063 | 219508064 | G | A | ENSG00000115568 | Brain - Cortex | Brain | -2.671205 | 0.47720839 |
| chr2 | 219508063 | 219508064 | G | A | ENSG00000115568 | Brain - Frontal Cortex (BA9) | Brain | -2.7664803 | 0.48028874 |
| chr2 | 219508063 | 219508064 | G | A | ENSG00000115568 | Brain - Hippocampus | Brain | -3.3785098 | 0.50010194 |
| chr2 | 219508063 | 219508064 | G | A | ENSG00000115568 | Brain - Hypothalamus | Brain | -2.7681826 | 0.48034379 |
| chr2 | 219508063 | 219508064 | G | A | ENSG00000115568 | Brain - Nucleus accumbens (basal ganglia) | Brain | -2.6200048 | 0.47555374 |
| chr2 | 219508063 | 219508064 | G | A | ENSG00000115568 | Brain - Putamen (basal ganglia) | Brain | -3.1809081 | 0.49370202 |
| chr2 | 219508063 | 219508064 | G | A | ENSG00000115568 | Brain - Spinal cord (cervical c-1) | Brain | -2.3156533 | 0.46573021 |
| chr2 | 219508063 | 219508064 | G | A | ENSG00000115568 | Brain - Substantia nigra | Brain | -2.6542191 | 0.4766594 |
| chr3 | 72881631 | 72881632 | C | A | ENSG00000144736 | Brain - Cerebellar Hemisphere | Brain | -0.1849922 | 0.87907336 |
| chr3 | 72881631 | 72881632 | C | A | ENSG00000144736 | Brain - Frontal Cortex (BA9) | Brain | -0.2238248 | 0.87766522 |
| chr3 | 72881631 | 72881632 | C | A | ENSG00000144736 | Brain - Hippocampus | Brain | -0.1863109 | 0.87902577 |
| chr3 | 72881631 | 72881632 | C | A | ENSG00000144736 | Brain - Hypothalamus | Brain | -0.1863109 | 0.87902577 |
| chr3 | 72881631 | 72881632 | C | A | ENSG00000144736 | Brain - Spinal cord (cervical c-1) | Brain | -0.1863109 | 0.87902577 |
| chr3 | 72881631 | 72881632 | C | A | ENSG00000144736 | Brain - Substantia nigra | Brain | -0.1863109 | 0.87902577 |
| chr3 | 72893597 | 72893598 | T | C | ENSG00000144736 | Brain - Cerebellar Hemisphere | Brain | -0.0633094 | 0.88339554 |
| chr3 | 72893597 | 72893598 | T | C | ENSG00000144736 | Brain - Frontal Cortex (BA9) | Brain | -0.1282273 | 0.88110659 |
| chr3 | 72893597 | 72893598 | T | C | ENSG00000144736 | Brain - Hippocampus | Brain | -0.0646281 | 0.88334942 |
| chr3 | 72893597 | 72893598 | T | C | ENSG00000144736 | Brain - Hypothalamus | Brain | -0.0646281 | 0.88334942 |
| chr3 | 72893597 | 72893598 | T | C | ENSG00000144736 | Brain - Spinal cord (cervical c-1) | Brain | -0.0646281 | 0.88334942 |

| | | | | | | | | | |
|---|---|---|---|---|---|---|---|---|---|
| chr3 | 72893597 | 72893598 | T | C | ENSG00000144736 | Brain - Substantia nigra | Brain | -0.0646281 | 0.88334942 |
| chr3 | 186505036 | 186505039 | TCA | T | ENSG00000156976 | Brain - Caudate (basal ganglia) | Brain | -1.0124405 | 0.07144223 |
| chr3 | 186505036 | 186505039 | TCA | T | ENSG00000156976 | Brain - Cerebellar Hemisphere | Brain | -1.0551993 | 0.07332787 |
| chr3 | 186505036 | 186505039 | TCA | T | ENSG00000156976 | Brain - Cortex | Brain | -1.0438588 | 0.07282335 |
| chr3 | 186505036 | 186505039 | TCA | T | ENSG00000156976 | Brain - Frontal Cortex (BA9) | Brain | -1.0335593 | 0.0723679 |
| chr3 | 186505036 | 186505039 | TCA | T | ENSG00000156976 | Brain - Hippocampus | Brain | -1.2083797 | 0.08046479 |
| chr3 | 186505036 | 186505039 | TCA | T | ENSG00000156976 | Brain - Hypothalamus | Brain | -1.3874482 | 0.08960766 |
| chr3 | 186505036 | 186505039 | TCA | T | ENSG00000156976 | Brain - Nucleus accumbens (basal ganglia) | Brain | -1.2651689 | 0.08326814 |
| chr3 | 186505036 | 186505039 | TCA | T | ENSG00000156976 | Brain - Putamen (basal ganglia) | Brain | -1.0480786 | 0.07301071 |
| chr3 | 186505036 | 186505039 | TCA | T | ENSG00000156976 | Brain - Spinal cord (cervical c-1) | Brain | -1.0480786 | 0.07301071 |
| chr3 | 186505036 | 186505039 | TCA | T | ENSG00000156976 | Brain - Substantia nigra | Brain | -1.0480786 | 0.07301071 |
| chr6 | 86256801 | 86256802 | T | C | ENSG00000135317 | Brain - Caudate (basal ganglia) | Brain | -0.5750492 | 0.99997611 |
| chr6 | 86256801 | 86256802 | T | C | ENSG00000135317 | Brain - Cerebellar Hemisphere | Brain | -0.4199903 | 0.99998302 |
| chr6 | 86256801 | 86256802 | T | C | ENSG00000135317 | Brain - Cortex | Brain | -0.5750492 | 0.99997611 |
| chr6 | 86256801 | 86256802 | T | C | ENSG00000135317 | Brain - Frontal Cortex (BA9) | Brain | -0.4199903 | 0.99998302 |
| chr6 | 86256801 | 86256802 | T | C | ENSG00000135317 | Brain - Hippocampus | Brain | -0.2730694 | 0.99998771 |
| chr6 | 86256801 | 86256802 | T | C | ENSG00000135317 | Brain - Hypothalamus | Brain | -0.5750492 | 0.99997611 |
| chr6 | 86256801 | 86256802 | T | C | ENSG00000135317 | Brain - Nucleus accumbens (basal ganglia) | Brain | -0.5750492 | 0.99997611 |
| chr6 | 86256801 | 86256802 | T | C | ENSG00000135317 | Brain - Putamen (basal ganglia) | Brain | -0.4199903 | 0.99998302 |
| chr6 | 86256801 | 86256802 | T | C | ENSG00000135317 | Brain - Spinal cord (cervical c-1) | Brain | -0.5750492 | 0.99997611 |
| chr6 | 86256801 | 86256802 | T | C | ENSG00000135317 | Brain - Substantia nigra | Brain | -0.5750492 | 0.99997611 |
| chr9 | 88252526 | 88252527 | G | C | ENSG00000135049 | Brain - Caudate (basal ganglia) | Brain | -0.0679415 | 0.99577247 |
| chr9 | 88252526 | 88252527 | G | C | ENSG00000135049 | Brain - Cerebellar Hemisphere | Brain | -0.0688679 | 0.99576858 |
| chr9 | 88252526 | 88252527 | G | C | ENSG00000135049 | Brain - Cortex | Brain | -0.0675492 | 0.99577412 |
| chr9 | 88252526 | 88252527 | G | C | ENSG00000135049 | Brain - Frontal Cortex (BA9) | Brain | -0.0675492 | 0.99577412 |
| chr9 | 88252526 | 88252527 | G | C | ENSG00000135049 | Brain - Hippocampus | Brain | -0.0681796 | 0.99577147 |

| | | | | | | | | | |
|---|---|---|---|---|---|---|---|---|---|
| chr9 | 88252526 | 88252527 | G | C | ENSG00000135049 | Brain - Hypothalamus | Brain | -0.0681796 | 0.99577147 |
| chr9 | 88252526 | 88252527 | G | C | ENSG00000135049 | Brain - Nucleus accumbens (basal ganglia) | Brain | -0.0688679 | 0.99576858 |
| chr9 | 88252526 | 88252527 | G | C | ENSG00000135049 | Brain - Putamen (basal ganglia) | Brain | -0.0681796 | 0.99577147 |
| chr9 | 88252526 | 88252527 | G | C | ENSG00000135049 | Brain - Spinal cord (cervical c-1) | Brain | -0.0675492 | 0.99577412 |
| chr9 | 88252526 | 88252527 | G | C | ENSG00000135049 | Brain - Substantia nigra | Brain | -0.0681796 | 0.99577147 |
| chr10 | 79769438 | 79769439 | G | C | ENSG00000148606 | Brain - Caudate (basal ganglia) | Brain | -0.0309617 | 0.93029199 |
| chr10 | 79769438 | 79769439 | G | C | ENSG00000148606 | Brain - Cerebellar Hemisphere | Brain | -0.0246999 | 0.93047242 |
| chr10 | 79769438 | 79769439 | G | C | ENSG00000148606 | Brain - Cortex | Brain | -0.0246999 | 0.93047242 |
| chr10 | 79769438 | 79769439 | G | C | ENSG00000148606 | Brain - Frontal Cortex (BA9) | Brain | -0.0246999 | 0.93047242 |
| chr10 | 79769438 | 79769439 | G | C | ENSG00000148606 | Brain - Hippocampus | Brain | -0.0397982 | 0.93003663 |
| chr10 | 79769438 | 79769439 | G | C | ENSG00000148606 | Brain - Hypothalamus | Brain | -0.0411169 | 0.92999845 |
| chr10 | 79769438 | 79769439 | G | C | ENSG00000148606 | Brain - Nucleus accumbens (basal ganglia) | Brain | -0.0411169 | 0.92999845 |
| chr10 | 79769438 | 79769439 | G | C | ENSG00000148606 | Brain - Putamen (basal ganglia) | Brain | -0.0387328 | 0.93006747 |
| chr10 | 79769438 | 79769439 | G | C | ENSG00000148606 | Brain - Spinal cord (cervical c-1) | Brain | -0.0399304 | 0.9300328 |
| chr10 | 79769438 | 79769439 | G | C | ENSG00000148606 | Brain - Substantia nigra | Brain | -0.0397982 | 0.93003663 |
| chr11 | 108144273 | 108144274 | G | T | ENSG00000149311 | Brain - Caudate (basal ganglia) | Brain | -0.0061888 | 0.99255135 |
| chr11 | 108144273 | 108144274 | G | T | ENSG00000149311 | Brain - Cerebellar Hemisphere | Brain | -0.0061888 | 0.99255135 |
| chr11 | 108144273 | 108144274 | G | T | ENSG00000149311 | Brain - Cortex | Brain | -0.0048701 | 0.9925599 |
| chr11 | 108144273 | 108144274 | G | T | ENSG00000149311 | Brain - Frontal Cortex (BA9) | Brain | -0.0061888 | 0.99255135 |
| chr11 | 108144273 | 108144274 | G | T | ENSG00000149311 | Brain - Hippocampus | Brain | -0.0055005 | 0.99255581 |
| chr11 | 108144273 | 108144274 | G | T | ENSG00000149311 | Brain - Hypothalamus | Brain | -0.0061888 | 0.99255135 |
| chr11 | 108144273 | 108144274 | G | T | ENSG00000149311 | Brain - Nucleus accumbens (basal ganglia) | Brain | -0.0061888 | 0.99255135 |
| chr11 | 108144273 | 108144274 | G | T | ENSG00000149311 | Brain - Putamen (basal ganglia) | Brain | -0.0048701 | 0.9925599 |
| chr11 | 108144273 | 108144274 | G | T | ENSG00000149311 | Brain - Spinal cord (cervical c-1) | Brain | -0.0050023 | 0.99255904 |
| chr11 | 108144273 | 108144274 | G | T | ENSG00000149311 | Brain - Substantia nigra | Brain | -0.0055005 | 0.99255581 |
| chr11 | 108144277 | 108144278 | A | C | ENSG00000149311 | Brain - Caudate (basal ganglia) | Brain | -0.0061888 | 0.99255135 |

| | | | | | | | | | |
|---|---|---|---|---|---|---|---|---|---|
| chr11 | 108144277 | 108144278 | A | C | ENSG00000149311 | Brain - Cerebellar Hemisphere | Brain | -0.0061888 | 0.99255135 |
| chr11 | 108144277 | 108144278 | A | C | ENSG00000149311 | Brain - Cortex | Brain | -0.0048701 | 0.9925599 |
| chr11 | 108144277 | 108144278 | A | C | ENSG00000149311 | Brain - Frontal Cortex (BA9) | Brain | -0.0061888 | 0.99255135 |
| chr11 | 108144277 | 108144278 | A | C | ENSG00000149311 | Brain - Hippocampus | Brain | -0.0055005 | 0.99255581 |
| chr11 | 108144277 | 108144278 | A | C | ENSG00000149311 | Brain - Hypothalamus | Brain | -0.0061888 | 0.99255135 |
| chr11 | 108144277 | 108144278 | A | C | ENSG00000149311 | Brain - Nucleus accumbens (basal ganglia) | Brain | -0.0061888 | 0.99255135 |
| chr11 | 108144277 | 108144278 | A | C | ENSG00000149311 | Brain - Putamen (basal ganglia) | Brain | -0.0048701 | 0.9925599 |
| chr11 | 108144277 | 108144278 | A | C | ENSG00000149311 | Brain - Spinal cord (cervical c-1) | Brain | -0.0050023 | 0.99255904 |
| chr11 | 108144277 | 108144278 | A | C | ENSG00000149311 | Brain - Substantia nigra | Brain | -0.0055005 | 0.99255581 |
| chr12 | 57036240 | 57036241 | C | A | ENSG00000110955 | Brain - Caudate (basal ganglia) | Brain | -2.0111654 | 0.98338017 |
| chr12 | 57036240 | 57036241 | C | A | ENSG00000110955 | Brain - Cerebellar Hemisphere | Brain | -1.8934632 | 0.98548117 |
| chr12 | 57036240 | 57036241 | C | A | ENSG00000110955 | Brain - Cortex | Brain | -1.8066246 | 0.98686133 |
| chr12 | 57036240 | 57036241 | C | A | ENSG00000110955 | Brain - Frontal Cortex (BA9) | Brain | -1.8934632 | 0.98548117 |
| chr12 | 57036240 | 57036241 | C | A | ENSG00000110955 | Brain - Hippocampus | Brain | -1.7203169 | 0.98810459 |
| chr12 | 57036240 | 57036241 | C | A | ENSG00000110955 | Brain - Hypothalamus | Brain | -1.9592438 | 0.9843415 |
| chr12 | 57036240 | 57036241 | C | A | ENSG00000110955 | Brain - Nucleus accumbens (basal ganglia) | Brain | -2.0111654 | 0.98338017 |
| chr12 | 57036240 | 57036241 | C | A | ENSG00000110955 | Brain - Putamen (basal ganglia) | Brain | -1.7732249 | 0.98735697 |
| chr12 | 57036240 | 57036241 | C | A | ENSG00000110955 | Brain - Spinal cord (cervical c-1) | Brain | -1.9488052 | 0.98452806 |
| chr12 | 57036240 | 57036241 | C | A | ENSG00000110955 | Brain - Substantia nigra | Brain | -1.7796156 | 0.98726358 |
| chr15 | 44898316 | 44898317 | T | C | ENSG00000104133 | Brain - Caudate (basal ganglia) | Brain | 0.04196113 | 0.99998856 |
| chr15 | 44898316 | 44898317 | T | C | ENSG00000104133 | Brain - Cerebellar Hemisphere | Brain | -0.5064238 | 0.99996411 |
| chr15 | 44898316 | 44898317 | T | C | ENSG00000104133 | Brain - Cortex | Brain | 0.04196113 | 0.99998856 |
| chr15 | 44898316 | 44898317 | T | C | ENSG00000104133 | Brain - Frontal Cortex (BA9) | Brain | -0.0598465 | 0.99998585 |
| chr15 | 44898316 | 44898317 | T | C | ENSG00000104133 | Brain - Hippocampus | Brain | 0.04196113 | 0.99998856 |
| chr15 | 44898316 | 44898317 | T | C | ENSG00000104133 | Brain - Hypothalamus | Brain | 0.04196113 | 0.99998856 |
| chr15 | 44898316 | 44898317 | T | C | ENSG00000104133 | Brain - Nucleus accumbens (basal ganglia) | Brain | -0.0598465 | 0.99998585 |

| | | | | | | | | | |
|---|---|---|---|---|---|---|---|---|---|
| chr15 | 44898316 | 44898317 | T | C | ENSG00000104133 | Brain - Putamen (basal ganglia) | Brain | -0.0598465 | 0.99998585 |
| chr15 | 44898316 | 44898317 | T | C | ENSG00000104133 | Brain - Spinal cord (cervical c-1) | Brain | 0.04196113 | 0.99998856 |
| chr15 | 44898316 | 44898317 | T | C | ENSG00000104133 | Brain - Substantia nigra | Brain | 0.04196113 | 0.99998856 |
| chr15 | 44943909 | 44943910 | G | C | ENSG00000104133 | Brain - Caudate (basal ganglia) | Brain | -0.973936 | 0.99990488 |
| chr15 | 44943909 | 44943910 | G | C | ENSG00000104133 | Brain - Cerebellar Hemisphere | Brain | -0.973936 | 0.99990488 |
| chr15 | 44943909 | 44943910 | G | C | ENSG00000104133 | Brain - Cortex | Brain | -0.973936 | 0.99990488 |
| chr15 | 44943909 | 44943910 | G | C | ENSG00000104133 | Brain - Frontal Cortex (BA9) | Brain | -0.973936 | 0.99990488 |
| chr15 | 44943909 | 44943910 | G | C | ENSG00000104133 | Brain - Hippocampus | Brain | -0.973936 | 0.99990488 |
| chr15 | 44943909 | 44943910 | G | C | ENSG00000104133 | Brain - Hypothalamus | Brain | -0.973936 | 0.99990488 |
| chr15 | 44943909 | 44943910 | G | C | ENSG00000104133 | Brain - Nucleus accumbens (basal ganglia) | Brain | -0.973936 | 0.99990488 |
| chr15 | 44943909 | 44943910 | G | C | ENSG00000104133 | Brain - Putamen (basal ganglia) | Brain | -0.973936 | 0.99990488 |
| chr15 | 44943909 | 44943910 | G | C | ENSG00000104133 | Brain - Spinal cord (cervical c-1) | Brain | -0.973936 | 0.99990488 |
| chr15 | 44943909 | 44943910 | G | C | ENSG00000104133 | Brain - Substantia nigra | Brain | -0.973936 | 0.99990488 |
| chr17 | 7796887 | 7796888 | G | A | ENSG00000170004 | Brain - Caudate (basal ganglia) | Brain | -0.1271223 | 0.13864794 |
| chr17 | 7796887 | 7796888 | G | A | ENSG00000170004 | Brain - Cerebellar Hemisphere | Brain | 0.27743361 | 0.12036288 |
| chr17 | 7796887 | 7796888 | G | A | ENSG00000170004 | Brain - Cortex | Brain | -0.9909696 | 0.18546925 |
| chr17 | 7796887 | 7796888 | G | A | ENSG00000170004 | Brain - Frontal Cortex (BA9) | Brain | -0.8266733 | 0.17570972 |
| chr17 | 7796887 | 7796888 | G | A | ENSG00000170004 | Brain - Hippocampus | Brain | -0.1066728 | 0.13767029 |
| chr17 | 7796887 | 7796888 | G | A | ENSG00000170004 | Brain - Hypothalamus | Brain | -0.2453936 | 0.14441696 |
| chr17 | 7796887 | 7796888 | G | A | ENSG00000170004 | Brain - Nucleus accumbens (basal ganglia) | Brain | -0.014735 | 0.1333463 |
| chr17 | 7796887 | 7796888 | G | A | ENSG00000170004 | Brain - Putamen (basal ganglia) | Brain | -0.1133886 | 0.13799072 |
| chr17 | 7796887 | 7796888 | G | A | ENSG00000170004 | Brain - Spinal cord (cervical c-1) | Brain | -0.0767163 | 0.13624859 |
| chr17 | 7796887 | 7796888 | G | A | ENSG00000170004 | Brain - Substantia nigra | Brain | -0.1227029 | 0.13843616 |
| chr18 | 43666102 | 43666104 | CA | C | ENSG00000152234 | Brain - Caudate (basal ganglia) | Brain | -3.0394813 | 0.8838417 |
| chr18 | 43666102 | 43666104 | CA | C | ENSG00000152234 | Brain - Cerebellar Hemisphere | Brain | -3.3972784 | 0.85157828 |
| chr18 | 43666102 | 43666104 | CA | C | ENSG00000152234 | Brain - Cortex | Brain | -3.308043 | 0.86025837 |

| | | | | | | | | | |
|---|---|---|---|---|---|---|---|---|---|
| chr18 | 43666102 | 43666104 | CA | C | ENSG00000152234 | Brain - Frontal Cortex (BA9) | Brain | -3.0394813 | 0.8838417 |
| chr18 | 43666102 | 43666104 | CA | C | ENSG00000152234 | Brain - Hippocampus | Brain | -2.660154 | 0.91121822 |
| chr18 | 43666102 | 43666104 | CA | C | ENSG00000152234 | Brain - Hypothalamus | Brain | -2.6416618 | 0.91239146 |
| chr18 | 43666102 | 43666104 | CA | C | ENSG00000152234 | Brain - Nucleus accumbens (basal ganglia) | Brain | -3.0394813 | 0.8838417 |
| chr18 | 43666102 | 43666104 | CA | C | ENSG00000152234 | Brain - Putamen (basal ganglia) | Brain | -2.8406742 | 0.89900126 |
| chr18 | 43666102 | 43666104 | CA | C | ENSG00000152234 | Brain - Spinal cord (cervical c-1) | Brain | -3.0394813 | 0.8838417 |
| chr18 | 43666102 | 43666104 | CA | C | ENSG00000152234 | Brain - Substantia nigra | Brain | -2.3418588 | 0.9295453 |
| chr20 | 2845684 | 2845685 | A | AG | ENSG00000215305 | Brain - Caudate (basal ganglia) | Brain | -2.4216598 | 0.3079829 |
| chr20 | 2845684 | 2845685 | A | AG | ENSG00000215305 | Brain - Cerebellar Hemisphere | Brain | -2.4216598 | 0.3079829 |
| chr20 | 2845684 | 2845685 | A | AG | ENSG00000215305 | Brain - Cortex | Brain | -2.4188755 | 0.30776445 |
| chr20 | 2845684 | 2845685 | A | AG | ENSG00000215305 | Brain - Frontal Cortex (BA9) | Brain | -2.5493245 | 0.31809091 |
| chr20 | 2845684 | 2845685 | A | AG | ENSG00000215305 | Brain - Hippocampus | Brain | -2.3069544 | 0.29905499 |
| chr20 | 2845684 | 2845685 | A | AG | ENSG00000215305 | Brain - Hypothalamus | Brain | -2.4216598 | 0.3079829 |
| chr20 | 2845684 | 2845685 | A | AG | ENSG00000215305 | Brain - Nucleus accumbens (basal ganglia) | Brain | -2.3669026 | 0.30370249 |
| chr20 | 2845684 | 2845685 | A | AG | ENSG00000215305 | Brain - Putamen (basal ganglia) | Brain | -2.0496773 | 0.27958247 |
| chr20 | 2845684 | 2845685 | A | AG | ENSG00000215305 | Brain - Spinal cord (cervical c-1) | Brain | -2.3617116 | 0.30329844 |
| chr20 | 2845684 | 2845685 | A | AG | ENSG00000215305 | Brain - Substantia nigra | Brain | -2.3406011 | 0.30165844 |
| chr20 | 2841437 | 2841438 | C | T | ENSG00000215305 | Brain - Caudate (basal ganglia) | Brain | -2.5511674 | 0.36090352 |
| chr20 | 2841437 | 2841438 | C | T | ENSG00000215305 | Brain - Cerebellar Hemisphere | Brain | -2.5600501 | 0.361516 |
| chr20 | 2841437 | 2841438 | C | T | ENSG00000215305 | Brain - Cortex | Brain | -2.5657891 | 0.36191195 |
| chr20 | 2841437 | 2841438 | C | T | ENSG00000215305 | Brain - Frontal Cortex (BA9) | Brain | -2.5657891 | 0.36191195 |
| chr20 | 2841437 | 2841438 | C | T | ENSG00000215305 | Brain - Hippocampus | Brain | -2.5569064 | 0.36129918 |
| chr20 | 2841437 | 2841438 | C | T | ENSG00000215305 | Brain - Hypothalamus | Brain | -2.5511674 | 0.36090352 |
| chr20 | 2841437 | 2841438 | C | T | ENSG00000215305 | Brain - Nucleus accumbens (basal ganglia) | Brain | -2.5569064 | 0.36129918 |
| chr20 | 2841437 | 2841438 | C | T | ENSG00000215305 | Brain - Putamen (basal ganglia) | Brain | -2.0199462 | 0.32515267 |
| chr20 | 2841437 | 2841438 | C | T | ENSG00000215305 | Brain - Spinal cord (cervical c-1) | Brain | -2.5511674 | 0.36090352 |

| | | | | | | | | | |
|---|---|---|---|---|---|---|---|---|---|
| chr20 | 2841437 | 2841438 | C | T | ENSG00000215305 | Brain - Substantia nigra | Brain | -2.3874208 | 0.34969553 |
| chr20 | 2841437 | 2841438 | C | T | ENSG00000215305 | Brain - Caudate (basal ganglia) | Brain | -2.5511674 | 0.34352627 |
| chr20 | 2841437 | 2841438 | C | T | ENSG00000215305 | Brain - Cerebellar Hemisphere | Brain | -2.5600501 | 0.34418078 |
| chr20 | 2841437 | 2841438 | C | T | ENSG00000215305 | Brain - Cortex | Brain | -2.5657891 | 0.34460396 |
| chr20 | 2841437 | 2841438 | C | T | ENSG00000215305 | Brain - Frontal Cortex (BA9) | Brain | -2.5657891 | 0.34460396 |
| chr20 | 2841437 | 2841438 | C | T | ENSG00000215305 | Brain - Hippocampus | Brain | -2.5569064 | 0.34394907 |
| chr20 | 2841437 | 2841438 | C | T | ENSG00000215305 | Brain - Hypothalamus | Brain | -2.5511674 | 0.34352627 |
| chr20 | 2841437 | 2841438 | C | T | ENSG00000215305 | Brain - Nucleus accumbens (basal ganglia) | Brain | -2.5569064 | 0.34394907 |
| chr20 | 2841437 | 2841438 | C | T | ENSG00000215305 | Brain - Putamen (basal ganglia) | Brain | -2.0199462 | 0.30552865 |
| chr20 | 2841437 | 2841438 | C | T | ENSG00000215305 | Brain - Spinal cord (cervical c-1) | Brain | -2.5511674 | 0.34352627 |
| chr20 | 2841437 | 2841438 | C | T | ENSG00000215305 | Brain - Substantia nigra | Brain | -2.3874208 | 0.33156925 |
| chr20 | 49194968 | 49194969 | C | T | ENSG00000196396 | Brain - Caudate (basal ganglia) | Brain | -4.2331915 | 0.69278956 |
| chr20 | 49194968 | 49194969 | C | T | ENSG00000196396 | Brain - Cerebellar Hemisphere | Brain | -4.816214 | 0.62332168 |
| chr20 | 49194968 | 49194969 | C | T | ENSG00000196396 | Brain - Cortex | Brain | -4.4451761 | 0.66833238 |
| chr20 | 49194968 | 49194969 | C | T | ENSG00000196396 | Brain - Frontal Cortex (BA9) | Brain | -4.7129842 | 0.63609962 |
| chr20 | 49194968 | 49194969 | C | T | ENSG00000196396 | Brain - Hippocampus | Brain | -2.9986451 | 0.81284517 |
| chr20 | 49194968 | 49194969 | C | T | ENSG00000196396 | Brain - Hypothalamus | Brain | -3.4355963 | 0.77497695 |
| chr20 | 49194968 | 49194969 | C | T | ENSG00000196396 | Brain - Nucleus accumbens (basal ganglia) | Brain | -4.6301967 | 0.64621121 |
| chr20 | 49194968 | 49194969 | C | T | ENSG00000196396 | Brain - Putamen (basal ganglia) | Brain | -3.7555046 | 0.74398667 |
| chr20 | 49194968 | 49194969 | C | T | ENSG00000196396 | Brain - Spinal cord (cervical c-1) | Brain | -2.6484791 | 0.83949896 |
| chr20 | 49194968 | 49194969 | C | T | ENSG00000196396 | Brain - Substantia nigra | Brain | -2.3446686 | 0.86006086 |
| chrX | 21887795 | 21887796 | G | A | ENSG00000012174 | Brain - Caudate (basal ganglia) | Brain | -0.5725068 | 0.6321225 |
| chrX | 21887795 | 21887796 | G | A | ENSG00000012174 | Brain - Cerebellar Hemisphere | Brain | -1.1403592 | 0.62390907 |
| chrX | 21887795 | 21887796 | G | A | ENSG00000012174 | Brain - Cortex | Brain | -0.6427706 | 0.63111024 |
| chrX | 21887795 | 21887796 | G | A | ENSG00000012174 | Brain - Frontal Cortex (BA9) | Brain | -0.4716242 | 0.63357384 |
| chrX | 21887795 | 21887796 | G | A | ENSG00000012174 | Brain - Hippocampus | Brain | -0.6355083 | 0.63121492 |

| | | | | | | | | | |
|---|---|---|---|---|---|---|---|---|---|
| chrX | 21887795 | 21887796 | G | A | ENSG00000012174 | Brain - Hypothalamus | Brain | -0.6427706 | 0.63111024 |
| chrX | 21887795 | 21887796 | G | A | ENSG00000012174 | Brain - Nucleus accumbens (basal ganglia) | Brain | -0.6427706 | 0.63111024 |
| chrX | 21887795 | 21887796 | G | A | ENSG00000012174 | Brain - Spinal cord (cervical c-1) | Brain | -0.574833 | 0.63208901 |
| chrX | 21887795 | 21887796 | G | A | ENSG00000012174 | Brain - Substantia nigra | Brain | -0.6427706 | 0.63111024 |
